# Comprehensive Comparison of Sixteen Markers of Biological Aging: Cross-Sectional and Longitudinal Results from the Berlin Aging Study II (BASE-II)

**DOI:** 10.1101/2025.04.09.25325514

**Authors:** Valentin Max Vetter, Johanna Drewelies, Jan Homann, Sandra Düzel, Laura Deecke, Philippe Jawinski, Simone Kühn, Elisa Kubala, Sebastian Markett, Michael Mülleder, Markus Ralser, Ulman Lindenberger, Christina M. Lill, Denis Gerstorf, Lars Bertram, Ilja Demuth

## Abstract

**Introduction:** The disproportionate increase in lifespan compared to health span over the past decades results in a growing proportion of life marked by diseases, even if incidence rates are falling in some cases. However, not everyone ages at the same pace and some people remain in good health and preserve physical and cognitive function into old age. To quantify inter-individual differences in the biological aging process, numerous indicators of biological age have been developed. While these markers have often been validated individually, comparisons in the same people are scarce, complicating their evaluation and translation into clinical practice.

**Methods:** In this study, we analyzed 16 measures of biological aging including epigenetic clocks, proteomics clock, telomere length, and SkinAge, laboratory composite markers (BioAge, Allostatic Load), psychological aging, and Brain Age. These age markers were evaluated cross-sectionally as well as longitudinally in the context of age-associated outcomes covering frailty, mobility, cognitive function, depressive symptoms, autonomy in daily life, nutrition, morbidity, and chronic disease in participants of the Berlin Aging Study II (BASE-II).

**Results:** Longitudinal data was available for 1,083 participants with a mean age of 68.3 years at baseline (52% women) and an average follow-up period of 7.4 years. Correlation among markers of aging from different domains was low (r≤0.31). Allostatic Load Index and DunedinPACE showed the strongest and most consistent cross-sectional and longitudinal associations with age-associated phenotypes, including morbidity, cardiovascular health, and frailty. Both biomarkers individually increased the accuracy of a logistic regression model trained to predict incident cases of Metabolic Syndrome, high cardiovascular risk (Lifes’s Simple 7) as well as incident frailty (Fried’s frailty index) 7.4 years after baseline examination by up to 24 percentage points.

**Conclusion:** Our findings support the previously shown distinction between indicators of aging and provide a comprehensive overview of their individual strengths and weaknesses in the context of wide variety of age-associated phenotypes. Furthermore, we show their distinct ability to predict aging-related adverse outcomes and suggest a potential use-case in longitudinal prediction modelling.

## Introduction

Advances in healthcare, hygiene, lifestyle, and housing have led to an increase in the average lifespan in many countries over the past decades (1). However, this expansion in lifespan was not matched by the increase in healthspan, the time spent before the onset of chronic disease or age-associated impairments (2–4). At the same time, individual aging trajectories vary, and this heterogeneity increases with advancing age. While some individuals maintain good physical and mental health into old age, others show an early onset of chronic disease and impairment (5). The geroscience hypothesis states that interventions targeting the biological process of aging may result in an increase of healthspan and prevent or at least postpone the onset of chronic disease (6). Therefore, interventions that slow down or even reverse biological aging processes are of high individual but also societal interest (7) but traditionally require long follow-up periods (3, 8). To test interventions in a cost- and time-effective manner, the validation of markers of aging that “either alone or in a composite predict biological age” (3) are needed. Although extensive efforts have been made to identify and develop markers that can quantify inter-individual differences in biological age, no consensus has been reached (5, 9–11). This might be partly due to the differences in the conceptual meaning and the underlying framework of aging they aim to quantify and the methodological approach used. For example, aging clocks based on “-omics” data, like the proteomic clock or the first-generation epigenetic clocks, were trained to predict chronological age and the difference between the predicted and the actual chronological age was shown to bear biological meaning (12). In contrast, more recent versions of the epigenetic clocks are trained to predict age- and mortality-dependent biological changes or the so-called Pace of Aging (13, 14), and thus differ fundamentally in their underlying conceptualization. Other markers, such as BioAge (15) and Allostatic Load (16), are composite markers that aggregate laboratory measures associated with multiple organ systems into one composite biomarker of aging. Consistent with findings from our and other groups is that different indicators of aging only show a low to moderate association with each other (5, 9, 17–25). This may result from the fact that the indicators were derived from different aging domains using varying methods. Whilst existing indicators of aging are individually well-evaluated, comparisons of different markers within the same study population and their relation to age-associated phenotypes are scarce (17, 23). Further, by focusing on selected health outcomes or high-risk populations, these studies limit our understanding of the broader predictive validity of aging markers in the general population.

To close this gap, we evaluated 16 markers of aging by investigating their relationship to a wide range of physical and mental age-associated health phenotypes in 1,083 participants of the Berlin Aging Study II (BASE-II) based on both cross-sectional and longitudinal data. To assess their potential for clinical application, we additionally investigated the ability of each indicator to predict functional, physical and cognitive impairment after a mean follow-up period of 7.4-years. By doing so, we aim to provide a comprehensive overview of their individual strengths and weaknesses, in order to sharpen each biomarker’s individual profile, and contribute to a roadmap for their effective and targeted use in research and ultimately in clinical practice.

## Methods

### Study population

BASE-II is an observational longitudinal study aiming at the identification of factors that predict and shape healthy aging trajectories (26). Apparently healthy participants were recruited through the Max Planck Institute for Human Development’s participant pool in Berlin, as well as through advertisements in local newspapers and public transportation networks. Participants between the ages of 60 and 80 years (older cohort) and 20 and 35 years (younger cohort, not analyzed in this study) were eligible for recruitment. Men and women were recruited in equal numbers. The baseline examination (T0) of 1,671 older participants (medical part) was conducted between 2009 and 2014 (26). A follow-up assessment (T1) as part of the GendAge study was conducted between 2018 and 2020 (27) on average 7.4 years after baseline (SD: 1.5 years, range: 3.9 to 10.4 years). Only participants who had provided information on outcomes of interest at baseline as well as at follow-up were included in the final sample (n=1,083, Supplementary Figure 1), to allow comparison of the aging markers in the same cohort of individuals. Of the 588 participants who dropped out between baseline and follow-up, 126 were confirmed to have died. Reasons for dropout among the remaining 462 participants were not systematically evaluated. Differences in the variables analyzed in this study between the main study sample and participants who dropped-out are small and we do not expect that loss to follow-up substantially altered our findings (Supplementary Table 1).

All participants gave written informed consent. All assessments at baseline and follow-up were conducted in accordance with the Declaration of Helsinki and approved by the Ethics Committee of the Charité—Universitätsmedizin Berlin (approval numbers EA2/029/09, EA2/144/16, and EA2/224/21) and were registered in the German Clinical Trials Registry as DRKS00009277 (BASE-II) and DRKS00016157 (GendAge). The Ethics Committee of the Max Planck Institute for Human Development approved the procedure, and the Ethics Committee of the German Society for Psychology (DGPs) additionally approved of the MRI protocol. This manuscript was created in accordance with the STROBE guidelines (28).

### Variables

The Horvath (29), Hannum (30), PhenoAge (31), GrimAge (32), and DunedinPACE (14) epigenetic clock algorithms were applied to derive DNA methylation age (DNAmAge) from methylation data measured with the “Infinium MethylationEPIC”, version 1, array (Illumina, Inc., USA). Methylation data for the 7-CpG epigenetic clock (18) was obtained through Single Nucleotide Prime Extension (SNuPE) (18, 33). DNAmA acceleration (DNAmAA) was calculated as unstandardized residuals of a leukocyte cell-count adjusted linear regression analysis of DNAmAge on chronological age. Proteomics Age was derived from 248 proteins measured by liquid chromatography-mass spectrometry (LC-MS). Proteomics age acceleration (ProteomicsAA) was calculated similarly to the DNAmAA as residuals from a linear regression of Proteomics Age on chronological age. Telomere length was assessed through quantitative real-time PCR (rLTL, (34)) as well as estimated by the algorithm proposed by Lu and colleagues (35) from methylation data (DNAmTL). The average results across three raters evaluating the number of lentigines from photos of the participants skin was used to derive SkinAge (15). BioAge is a composite score that aggregates 13 routine laboratory parameters that were identified for their association with mortality (15). The Allostatic Load Index (ALI) was computed by awarding a point to every participant within the high risk quartile of selected variables using the approach described by Seeman and colleagues (16). It additionally incorporates information about intake of relevant medication to include successfully treated and thereby masked dysregulation in its calculation (36, 37). Subjective Felt Age (SFA) was calculated as proportional discrepancy score from self-reported „felt age“ and chronological age (38). Subjective Life Expectancy (SLE) is calculated as the difference between the age participants expect to live to and their chronological age at the time of assessment. Subjective Health Expectancy (SHE) is the difference between the age participants expect to remain healthy and their chronological age at the time of assessment. Both SLE and SHE were adjusted for chronological age (39). In addition to these markers, BrainAge was available in a subgroup of n=255 BASE-II participants who underwent Magnetic Resonance Imaging (MRI) in a 3-Tesla Siemens Magnetom Trio scanner. BrainAge was calculated using a model trained in participants from the UK Biobank (40). Except for proteomics age, all other indicators of aging were individually investigated in BASE-II before (15, 18, 19, 41–49).

Outcome measures investigated in this study include two measures of frailty. The Fried Frailty Phenotype (Fried FI) includes information on unintended weight loss, exhaustion, weakness, slow walking speed, and low physical activity (50). The SPRINT-BASEed frailty index (SP FI) used in this study is an adapted version (51) of the deficit-based measure developed by Pajewsky and colleagues (52). Finger Floor Distance (FFD) was measured in centimeters. Mini Mental State Examination (MMSE) is a well-established interviewer-administered instrument which was used to assess cognitive impairment (53). Additionally, cognitive performance, specifically processing speed, was measured by Digit Symbol Substitution Test (DSST) for which participants were asked to match symbols to numbers according to a given key (54). Depressive symptoms were assessed using the Center for Epidemiologic Studies Depression Scale (CES-D) (55). Independence during everyday life and the ability to perform tasks without help was measured using the Activities of Daily Living questionnaire (ADL, “Barthel Index”) (56). The nutritional status was assessed using results from a short questionnaire and the measured circumference of the upper arm and the calf (Mini Nutritional Assessment, MNA (57)). Type 2 Diabetes (T2D) was diagnosed based on the criteria defined by the American Diabetes Association (ADA) guidelines (58). Diabetes associated complications were quantified using the composite score developed by Young and colleagues (59, 60). The Morbidity Index (MI) was calculated to assess the overall morbidity burden of the BASE-II participants by adapting the approach first described by Charlson and colleagues (61). The Systematic Coronary Risk Evaluation assessment (SCORE2 (62)) and the SCORE2-OP (for participants >70 years, hereafter referred to as SCORE2 (63)) were calculated according to the recommendations in the respective publications to assess the risk for cardiovascular events. An adapted version (41, 64) of the Life’s Simple 7 (LS7 (65)) was calculated to quantify modifiable cardiovascular risk factors. Metabolic Syndrome (MetS) was diagnosed using the definition suggested by the American Heart Association/ International Diabetes Federation/ National Heart, Lung, and Blood Institute criteria 2009 (66).

Confounding variables included in the regression models were chronological age (years), sex, alcohol consumption (g/d), and nicotine consumption (packyears), which were assessed during 1:1-interviews with trained study personnel. Body Mass Index (BMI) was calculated as kg/m^2^ using measurements from an electronic measuring station (seca 763, SECA, Germany)). Genetic ancestry was quantified using the first four components from a principal component analysis on genome-wide single nucleotide polymorphism genotyping data (67). Details on the variables investigated in this study are described in the Supplementary Material.

### Statistical analysis

Statistical analyses and visualizations were conducted using R software (version 3.2.2) (68). Missing values were imputed by multiple imputation using the mice package (for detailed information, please see the Supplementary Methods). BrainAge was available in a subgroup of 255 BASE-II participants. To facilitate comparison with all other markers, a separate multiple imputation procedure including all other variables was applied in this subsample. Descriptive statistics of the first imputed dataset are presented in Table 1 and 2. Descriptive statistics of the original (non-imputed) dataset are shown in Supplementary Table 2. Intercorrelation plots for markers of aging and outcome variables are presented in Supplementary Figure 2 and Supplementary Figure 3. The change in outcome variables was assessed by subtracting the outcome value at T0 from the value documented for T1. The relationship between markers of aging and outcome variables was assessed by logistic and linear regression models. In line with previous publications in the field as well as recommendations for increased quality in biomarker validation, we report results from an unadjusted, a minimally adjusted (chronological age and sex), and fully adjusted model (69). The longitudinal minimally and fully adjusted model is additionally adjusted for the outcome variables at T0. Since we were mainly interested in the raw predictive ability of the biomarkers (diagnostically as well as prognostically), we focus on the description of the unadjusted model in the main text. Statistical significance of the differences between the basic risk prediction models with and without the respective marker of aging was assessed using the *roc.*test-function (pROC package). To adjust for multiple testing, a Bonferroni correction was applied to the main analyses. As cross-sectional and longitudinal analyses were performed, we set our level for statistical significance at α = 0.05/(2*16 markers*16 outcomes) = 0.0001. To facilitate comparison between markers and outcome measure on differing scales, all values were z-transformed using R’s *scale* function prior to the regression analyses and standardized effect measures are reported. Dichotomization of continuously scaled variables used in the sensitivity analyses and the impairment prediction models was done using validated cut-off values if available (Supplementary Table 3). The MMSE and ADL were excluded from analyses that examined the dichotomized variables due to the very limited number of participants showing impairment according to these instruments. For the main analyses, sex-stratified subgroup analyses were performed as suggested by Moqri and colleagues (69). No age-group stratified analyses are shown due to the already narrow age range in BASE-II (69). Further information on the statistical analyses can be found in the supplementary material.

**Table 1:** Descriptive statistics of markers of aging at baseline in older participants of the BASE-II (n=1,083).

| Variables | Mean (SD) | Min | Max |
| --- | --- | --- | --- |
| 7-CpG DNAmAA | -0.02 (6.94) | -22.93 | 22.33 |
| Horvath DNAmAA | 0.08 (4.22) | -11.64 | 14.45 |
| Hannum DNAmAA | -0.04 (3.43) | -10.9 | 19.15 |
| PhenoAge DNAmAA | 0.08 (4.65) | -16.88 | 14.28 |
| GrimAge DNAmAA | 0.07 (3.12) | -8.52 | 10.2 |
| DunedinPACE | 1.01 (0.11) | 0.58 | 1.42 |
| DNAmTL | 6.97 (0.18) | 6.26 | 7.44 |
| rLTL | 1.15 (0.23) | 0.17 | 1.92 |
| SkinAge | 1.71 (0.90) | 0 | 3 |
| ProteomicsAA | -0.02 (1.34) | -4.18 | 5.11 |
| BioAge | 0.17 (5.91) | -16.37 | 20.21 |
| Allostatic Load | 3.98 (2.40) | 0 | 12 |
| Subj. Age* | 0.09 (0.08) | -0.13 | 0.41 |
| Subj. Life Expectancy | 15.89 (7.68) | 0 | 47 |
| Subj. Health Expectancy | 11.47 (7.06) | 0 | 46 |
| BrainAge** | -0.06 (3.23) | -9.03 | 8.03 |

Note: \* Subj. Age = relative subjective age adjusted by chronological age. \*\* BrainAge was available in a subgroup of 255 participants.

## Results

### Participants

In this study, 1,083 BASE-II participants who provided information at baseline and follow-up on average 7.4 years later were analyzed with respect to cross-sectional and longitudinal associations of 16 markers of aging (Supplementary Figure 4) with a range of age-associated outcomes. Mean chronological age at baseline was 68.3 years (SD: 3.5 years) and 52% were women (Table 1). Frequency of impairment in the analyzed variables is shown in Supplementary Table 4. All age-associated outcome variables differed statistically significantly between examinations with the exception of FFD, MMSE, ADL, and CES-D (Table 2). As described in BASE-II before (24), correlations between markers of aging from different domains were moderate to low with r≤0.31 in all instances (Supplementary Figure 2).

**Table 2:** Descriptive statistics of outcome variables at baseline (T0) and follow-up (T1) examination (n=1,083). Paired t-test and McNemar’s test were used to assess statistical significance of differences between timepoints.

| Variable | T0 |  |  |  | T1 |  |  |  | Mean Difference | SMD | p |
| --- | --- | --- | --- | --- | --- | --- | --- | --- | --- | --- | --- |
|  | Mean, n | SD, % | Min | Max | Mean, n | SD, % | Min | Max |  |  |  |
| Chronological Age | 68.272 | 3.489 | 60.159 | 84.632 | 75.62 | 3.768 | 64.912 | 94.073 | 7.348 | 2.024 | <0.001 |
| **Frailty and Mobility** |  |  |  |  |  |  |  |  |  |  |  |
| Fried Frailty | 0.364 | 0.61 | 0 | 3 | 0.763 | 0.878 | 0 | 4 | 0.399 | 0.528 | <0.001 |
| Frailty SPRINT-BASEed | 0.132 | 0.063 | 0.008 | 0.44 | 0.16 | 0.075 | 0 | 0.523 | 0.028 | 0.411 | <0.001 |
| Finger Floor Distance | 9.746 | 11.839 | 0 | 52 | 10.576 | 12.673 | 0 | 159 | 0.829 | 0.068 | 0.005 |
| Tinetti Score | 27.629 | 1.691 | 0 | 28 | 26.572 | 2.668 | 7 | 28 | -1.056 | -0.473 | <0.001 |
| Falls (past 12 months) | 336 | 0.31 |  |  | 295 | 0.272 |  |  |  |  | 0.043 |
| **Cognitive and Mental Health** |  |  |  |  |  |  |  |  |  |  |  |
| MMSE | 28.59 | 1.335 | 22 | 30 | 28.578 | 1.524 | 18 | 30 | -0.012 | -0.008 | 0.827 |
| DSST | 45.001 | 8.301 | 5 | 74 | 40.801 | 8.674 | 2 | 73 | -4.2 | -0.495 | <0.001 |
| CES-D | 7.285 | 6.613 | 0 | 36 | 7.17 | 6.165 | 0 | 36 | -0.115 | -0.018 | 0.526 |
| **Functional Assessments** |  |  |  |  |  |  |  |  |  |  |  |
| ADL | 99.003 | 3.72 | 5 | 100 | 98.698 | 4.086 | 15 | 100 | -0.305 | -0.078 | 0.046 |
| MNA | 27.383 | 1.756 | 19 | 30 | 26.595 | 2.158 | 16 | 30 | -0.788 | -0.401 | <0.001 |
| **Morbidity** |  |  |  |  |  |  |  |  |  |  |  |
| DCSI | 0.777 | 1.163 | 0 | 7 | 1.264 | 1.486 | 0 | 8 | 0.488 | 0.365 | <0.001 |
| Morbidity Index | 0.975 | 1.208 | 0 | 7 | 1.432 | 1.542 | 0 | 9 | 0.457 | 0.33 | <0.001 |
| SCORE2 | 10.809 | 4.222 | 4 | 32 | 16.073 | 6.16 | 5 | 47 | 5.264 | 0.997 | <0.001 |
| LS7 | 8.609 | 1.978 | 2 | 14 | 8.44 | 1.86 | 3 | 14 | -0.17 | -0.088 | 0.001 |
| T2D (diagnosed) | 127 | 0.117 |  |  | 185 | 0.171 |  |  |  |  | <0.001 |
| Metabolic Syndr. (diagnosed) | 388 | 0.358 |  |  | 491 | 0.453 |  |  |  |  | <0.001 |

Note: LS7 = Life's Simple Seven, SCORE2 = Systematic Coronary Risk Evaluation 2, MI = Charlson's Morbidity Index, DCSI = Diabetes complications severity index, MNA = Mini Nutritional Assessment, DSST = Digit Symbol Substitution Test, Tinetti = Tinetti Test, FFD = Finger Floor Distance, SP FI = SPRINT-BASEed Frailty Phenotype, Fried FI = Fried's Frailty Phenotype, MetS = Metabolic Syndrome, T2DM = Type 2 Diabetes Mellitus.

Cross-sectional association between markers of aging and age-associated outcomes at baseline Cross-sectional associations of markers of aging with continuously and categorically scaled age-associated outcomes at baseline were investigated using linear and logistic regression models. No statistically significant association (Bonferroni adjusted significance level defined at α = 0.0001) with the outcome variables was found for the first-generation epigenetic clocks, PhenoAge DNAmAA, SkinAge, Proteomics AA, and BrainAge. In contrast, GrimAge DNAmAA, DunedinPACE and both laboratory composite markers (BioAge, ALI) were statistically significantly associated with at least one of the two cardiovascular health scores (LS7, SCORE2, all p > 0.000018). In addition, ALI and DunedinPACE were associated with disease-related outcomes like MI (p=7.3×10^-6^), diagnosed T2D (p=5.7×10^-6^), and MetS (p=1.1×10^-10^, Supplementary Table 5). After correction for multiple testing, both TL measures were statistically significantly associated with FFD and SCORE2 and all three psychological aging markers (SFA, SLE, SHE) were statistically significantly associated with SPRINT-BASEed FI. Overall, the strongest and most robust results after covariate adjustment were found for ALI and DunedinPACE. All results from the unadjusted cross-sectional linear and logistic regression analyses as well as a minimally (age, sex) and fully adjusted models (age, sex, genetic ancestry, alcohol intake, smoking, BMI) are displayed in Figure 1, Supplementary Figure 5, and Supplementary Table 5 and 6.

**Figure 1:**
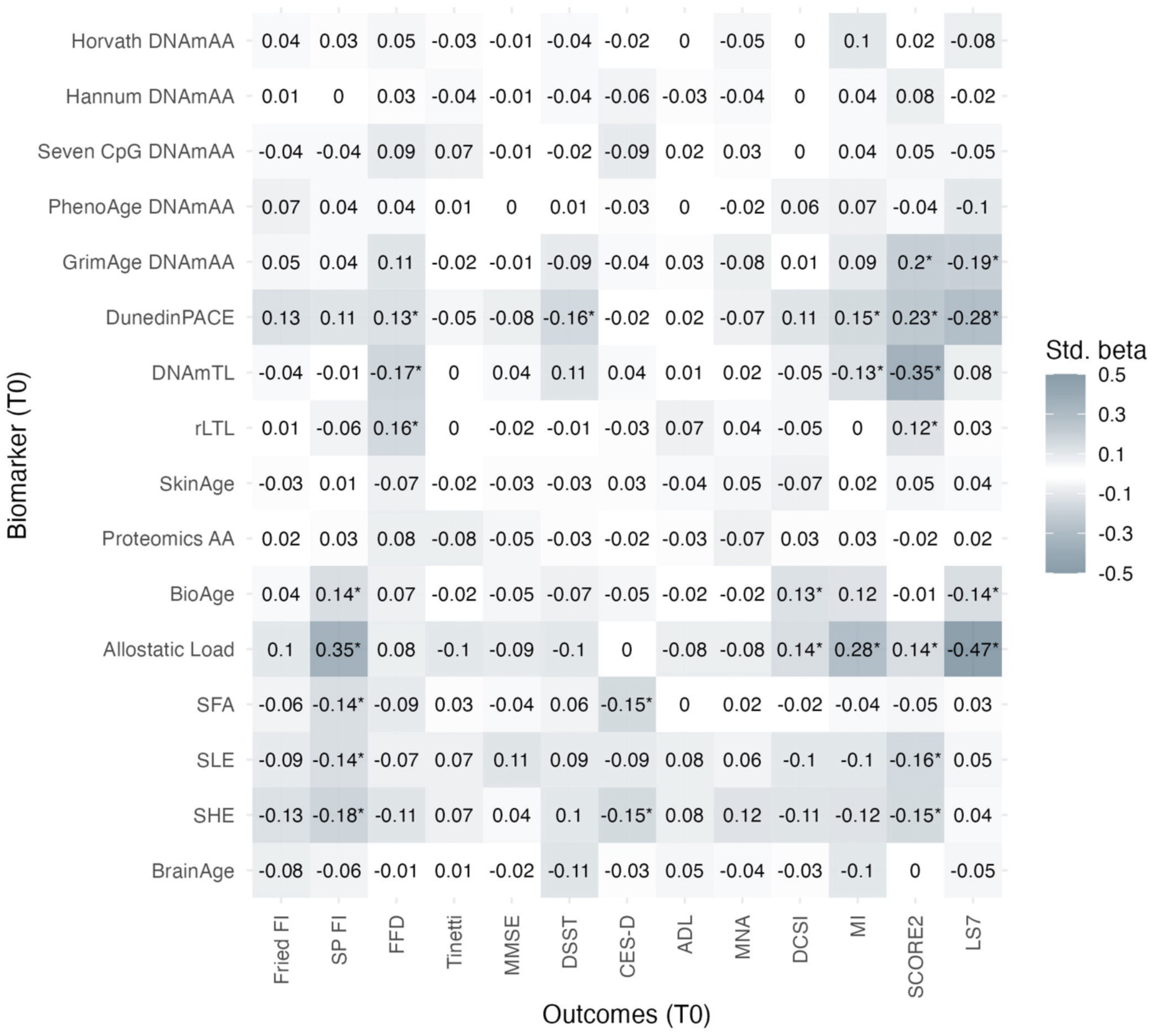
Standardized regression coefficients of unadjusted linear regression analyses of outcome variables on markers of aging in 1,083 participants from the BASE-II at baseline. BrainAge was available in a subgroup of n=255 participants. Note: * p <0.0001, LS7 = Life’s Simple Seven, SCORE2 = Systematic Coronary Risk Evaluation 2, MI = Charlson’s Morbidity Index, DCSI = Diabetes complications severity index, MNA = Mini Nutritional Assessment, DSST = Digit Symbol Substitution Test, Tinetti = Tinetti Test, FFD = Finger Floor Distance, SP FI = SPRINT-BASEed Frailty Phenotype, Fried FI = Fried’s Frailty Phenotype, MetS = Metabolic Syndrome, T2DM = Type 2 Diabetes Mellitus.

As a sensitivity analysis, we report on the results of logistic regression models of dichotomized continuously scaled variables on the markers of aging (Supplementary Table 6) and sex-stratified subgroup analyses (Supplementary Table 5 and 6). All analyses were repeated in the subsample of participants for which BrainAge was available (Supplementary Table 7, Supplementary Table 8) to allow direct comparison of results of BrainAge with the other markers in the same individuals.

Longitudinal associations between markers of aging at baseline and age-associated outcomes at 7.4 years of follow-up To explore the markers’ longitudinal association with the investigated outcomes, we performed regression analyses of outcomes measured at follow-up on markers assessed at baseline. Similarly to the cross-sectional analyses, no statistically significant associations with age-associated outcomes were found for measures derived from first-generation epigenetic clocks, PhenoAge, SkinAge, ProteomicsAA, and BrainAge. As already observed in the cross-sectional analyses, after correction for multiple testing the strongest and most consistent associations were found for DunedinPACE and the ALI. Interestingly, all three psychological aging markers were associated with Fried’s frailty phenotype in addition to the association with the SPRINT-BASEed frailty index that was already observed in the cross-sectional analyses at baseline (Figure 2, Supplementary Table 9). The longitudinal results of the dichotomized as well as of the binary outcome variables can be found in Supplementary Table 10 and Supplementary Figure 6. The results from the analyses of the subgroup of participants who provided BrainAge data can be found in Supplementary Table 11 and 12. Additionally, we investigated the change in the continuously scaled outcome variables by regressing the change in the outcome variables on the markers of aging at baseline which can be found in Supplementary Table 13. Similarly to the other longitudinal linear regression models, ALI showed the most frequent and consistent statistically significant associations across outcomes and models.

**Figure 2:**
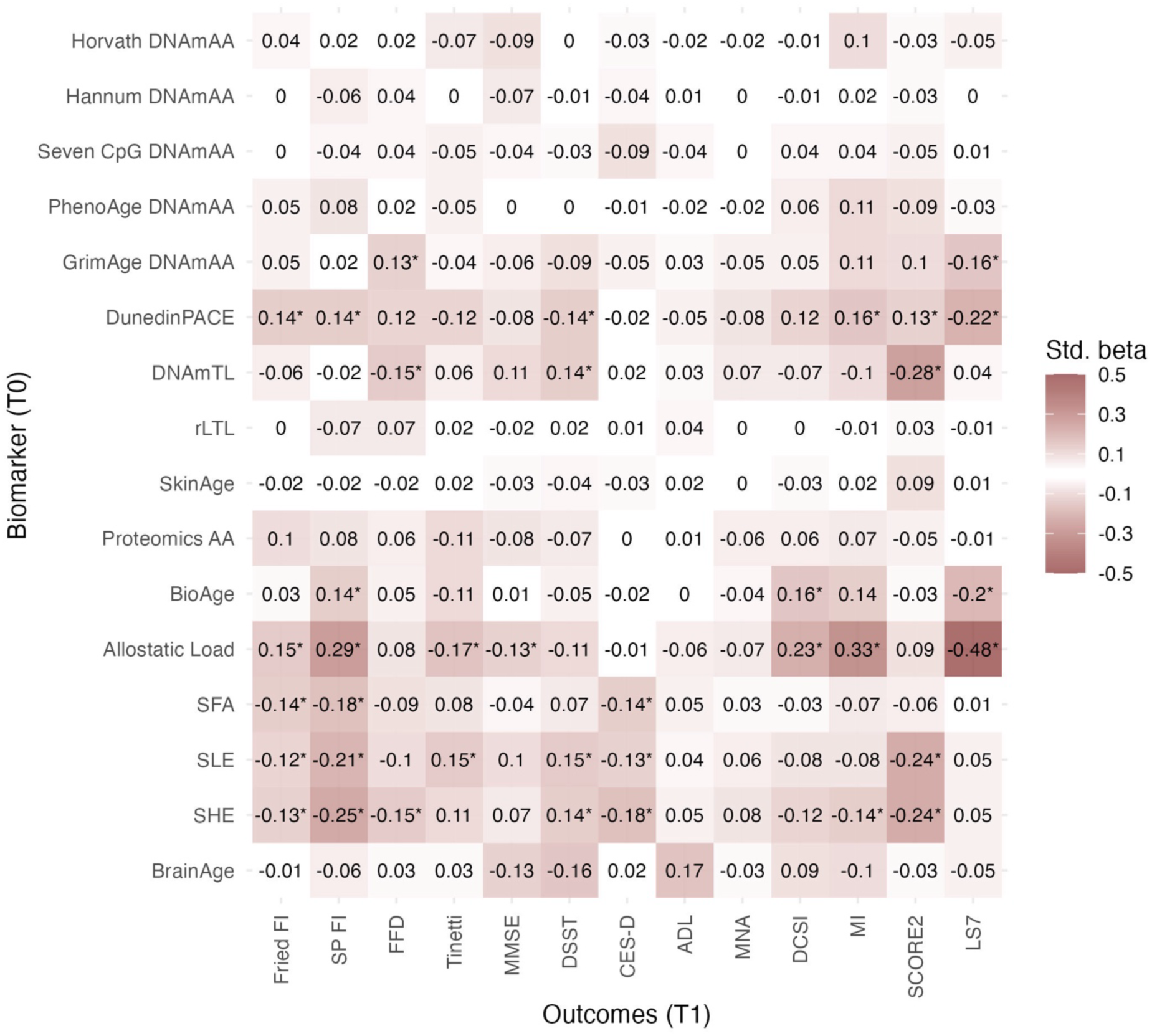
Standardized regression coefficients of unadjusted longitudinal linear regression analyses of outcome variables at T1 on markers of aging at T0 in 1,083 participants from the BASE-II. BrainAge was available in a subgroup of n=255 participants. Note: * p <0.0001, LS7 = Life’s Simple Seven, SCORE2 = Systematic Coronary Risk Evaluation 2, MI = Charlson’s Morbidity Index, DCSI = Diabetes complications severity index, MNA = Mini Nutritional Assessment, DSST = Digit Symbol Substitution Test, Tinetti = Tinetti Test, FFD = Finger Floor Distance, SP FI = SPRINT-BASEed Frailty Phenotype, Fried FI = Fried’s Frailty Phenotype, MetS = Metabolic Syndrome, T2DM = Type 2 Diabetes Mellitus.

### Prediction of incident cases in outcome variables over a 7.4-year follow-up period

To promote translation into clinical practice and to evaluate the markers in a possible use-case-scenario, we evaluated the markers’ ability to predict incidence of impairment in the analyzed assessments as well as incident cases of the investigated diseases and frailty over the average follow-up time of 7.4 years. Each marker-outcome-combination was evaluated in two scenarios. First, we simulated the case in which no other clinical information is available and assessed the markers raw predictive value. In a second step, we investigated the additional value that each marker adds to a basic prediction model.

Of all analyzed biomarkers, Allostatic Load was the only one to be statistically significantly (after multiple testing correction) associated with incident impairment at follow-up. Specifically, higher Allostatic Load was associated with increased odds for a diagnosis of T2D and MetS and its addition to the basic prediction model improved the AUC of the model by 24 and 18 percentage points (DeLong’s test, p<0.00001, Figure 4). Additionally, an increase in one SD of Allostatic Load at baseline increased the odds for the incidence of at least one diabetes-associated complication at T1 (DCSI) 1.4-times (p<0.0001) and the inclusion of Allostatic Load to a basic prediction model increased the AUC by 7 percentage points (p<0.00001). Odds for an incident classification as “average” or “inadequate” based on the LS7 criteria were 1.7-fold for every 1-SD increase in Allostatic Load (p= 0.00002, Figure 4) at baseline and it increased the accuracy of the basic prediction model by 20 percentage points (p<0.00001, Figure 4). Interestingly, while Allostatic Load was not statistically significantly associated with the outcome in prediction models for Fried’s frailty index and MI, its inclusion still improved the respective prediction models statistically significantly by 4 and 10 percentage points (p<0.00003), respectively. Similarly, DunedinPACE did not show statistically significant associations in the individual models, but its addition to the basic prediction model increased the AUC of models predicting diagnosis of MetS and impairment in Fried’s frailty index, and LS7 by 5, 4, and 8 percentage points (p<0.00006, Figure 4). In case of LS7, GrimAge DNAmAA improved the prediction model by 5 percentage points (p<0.0001, Figure 4). A very high AUC (AUC=0.935) was observed for the clinical prediction model with respect to identifying incident high cardiovascular risk defined by the SCORE2 algorithm. This prediction was significantly improved by 1 percentage point after adding TL measured using qPCR (p<0.00003). The complete results for all marker-outcome-combinations are shown in Figure 3 and Supplementary Table 14.

**Figure 3:**
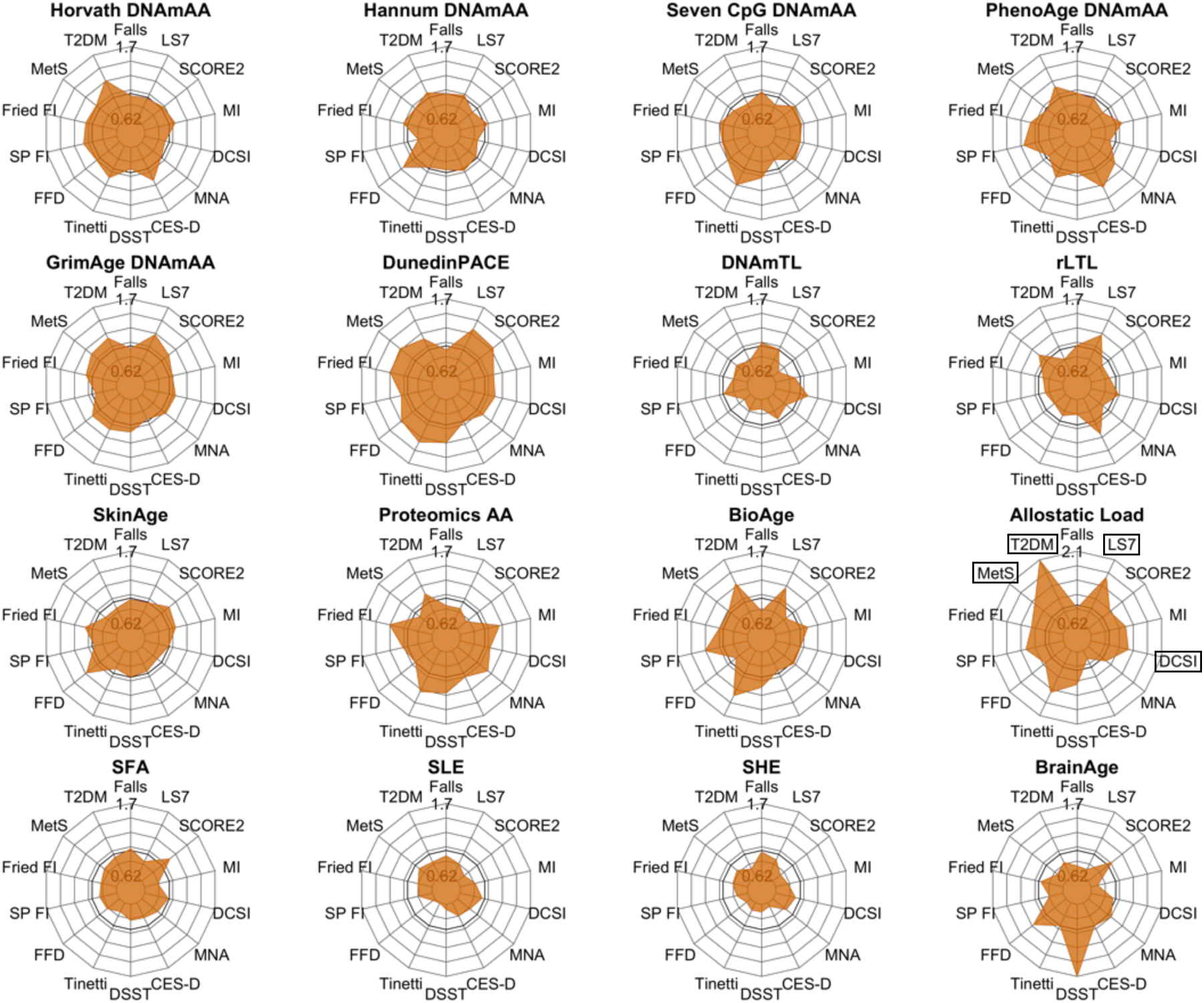
Radar plots showing the results of unadjusted logistic regression analyses of incident cases of analyzed outcome variables at T1 on markers at T0. Participants with prevalent cases at T0 were excluded from this analysis. Due to the multiple imputation procedure, the sample sizes between each imputed dataset differ slightly. The range of sample sizes are indicated in Supplementary Table 14. Due to very strong effect sizes observed for Allostatic Load the scale of this plot was adjusted to increase readability of the other plots. Therefore, the individual axis limits are displayed for each plot individually. Variable names of associations with p<0.0001 are displayed within a black box. Note: LS7 = Life’s Simple Seven, SCORE2 = Systematic Coronary Risk Evaluation 2, MI = Charlson’s Morbidity Index, DCSI = Diabetes complications severity index, MNA = Mini Nutritional Assessment, DSST = Digit Symbol Substitution Test, Tinetti = Tinetti Test, FFD = Finger Floor Distance, SP FI = SPRINT-BASEed Frailty Phenotype, Fried FI = Fried’s Frailty Phenotype, MetS = Metabolic Syndrome, T2DM = Type 2 Diabetes Mellitus.

**Figure 4:**
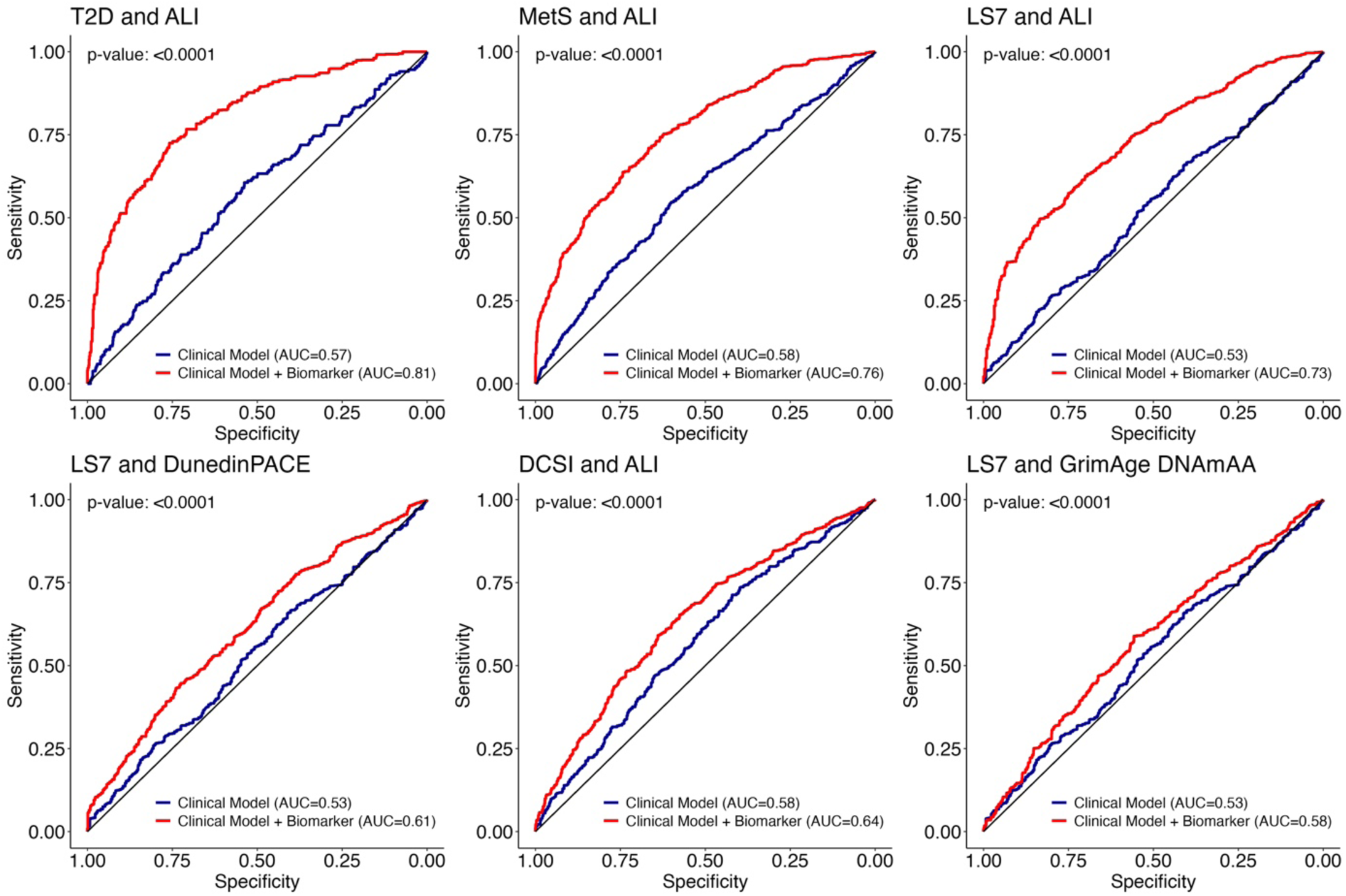
ROC curves illustrating the sensitivity and specificity of selected logistic regression models predicting impairment in age-associated phenotypes on average 7.4 years after marker assessment. The predictive performance of a basic clinical model including age and sex (blue) is shown compared to a prediction model which is extended by the respective markers of aging (red). The marker-outcome combinations presented here were selected to highlight the most compelling results from the three most promising markers (ALI, DunedinPACE, and GrimAge). A complete list of all AUC values can be found in Supplementary Table 14. P-values for the difference between prediction models were calculated using the approach described by DeLong (71) as part of the roc.test function (pROC package).

## Discussion

In this study, we investigated 16 markers of aging in 1,083 older participants from the Berlin Aging Study II (BASE-II). All markers were cross-sectionally and longitudinally analyzed in the context of a wide range of age-associated outcome variables representing different aspects of aging, including frailty, mobility, cognitive function, depressive symptoms, autonomy, nutrition, overall health, and chronic disease. Additionally, we investigated the markers’ ability to predict incident impairment in the age-associated outcomes over the average follow-up period of 7.4 years.

Our analyses showed that GrimAge DNAmAA and DunedinPACE performed best with respect to their association with cardiovascular health and cognitive capacity, Allostatic Load seems to represent frailty and overall morbidity associated variables (MI, T2D and MetS). Longitudinally, the subjective psychological markers were associated with depressive symptoms (CES-D) and cognitive functioning (DSST, except SFA). Additionally, they were associated with the SPRINT-BASEed frailty index (cross-sectionally and longitudinally) and Fried’s frailty index (longitudinally). The strongest effect sizes in cross-sectional and longitudinal analyses were found for ALI with respect to diagnosed T2D, MetS, LS7, MI and SPRINT-BASEed frailty index. This presumably results from the way it is calculated as many of the variables included in the ALI variable are closely related to health and are in some instances also part of the diagnostic criteria for T2D and MetS.

Subsequently, we investigated a potential use-case of the markers in a clinical context by predicting incident cases of impairment at follow-up by markers assessed on average 7.4-years earlier at baseline. ALI, GrimAge DNAmAA and DunedinPACE improved our basic prediction model in its prediction of incidence of impairment by up to 24 percentage points (Figure 4). As this increase in accuracy of the prediction model is expected to be less substantial when the markers are added to more comprehensive and specific prediction models, these results nevertheless illustrate that the markers hold biological information that can substantially improve the prediction of impairment years before it becomes clinically apparent.

Generally, we found more statistically significant results and stronger effect sizes in the longitudinal analyses compared to the cross-sectional results. This was observable especially for DunedinPACE, Allostatic Load and the psychological aging markers. Markers of aging are expected to depict the underlying aging processes which are anticipated to happen prior to clinical manifestations. Therefore, it is expected that an accelerated aging process that did not (yet) exceed the individual mechanisms of resilience and therefore did not manifest through clinical phenotypes would be trackable through markers. The results presented here suggest that the respective markers appear to recognize an acceleration in the underlying aging process that was (at least at baseline) not advanced enough or was still compensated by the physiological and cognitive coping mechanisms to not result in clinically observable phenotypes. However, at follow-up examination, the acceleration in the biological aging process, which was already picked up at baseline by the respective markers, potentially resulted in clinical manifestations. Another main finding of our study is the in part large differences in effect sizes of associations with the examined outcome variables which suggests that the analyzed markers capture distinct aspects of the biological aging processes (9, 17, 18, 25). This interpretation of our findings is particularly plausible due to the different concepts of biological age that underly the examined markers. For example, the first-generation epigenetic clocks aim at the prediction of chronological age and the residuals of a regression of chronological on epigenetic age are used to derive the analyzed marker (DNAmAA) (18, 29, 30). Second- and third-generation epigenetic clocks, in contrast, are trained on more complex measures of biological aging calculated from several individual variables (14, 31, 32) and DunedinPACE captures the rate of aging comparable to a speedometer (14, 70). Other markers, such as the psychological aging markers rely on the subjective self-assessment of the participants. Composite markers, like Allostatic Load and BioAge, on the other hand incorporate information from numerous systems to quantify aging while other measures, such as telomere length and BrainAge, aim at the quantification of age-related biological changes. These differences result in unique strengths and weaknesses of each marker which in turn define how they could potentially be used in the scientific and clinical context.

Our results are indicative that these markers are indeed capable of depicting underlying aging processes and do so with a higher sensitivity than clinical aging measures. Thereby, they could potentially be used to identify participants who are especially prone to future impairment due to an acceleration in one of the biological aging processes long before they show this decline clinically.

We want to point out several limitations to this study. Firstly, the participants of this study are above average health (26). Consequently, a comparatively low prevalence of impairment in the functional and cognitive assessments was observed which could lead to an underestimation of the true effect sizes. Secondly, the covariates used in the fully adjusted regression models were chosen because of their frequent use in other studies in the field as well as their known or suspected association with the independent and dependent variables in our regression models. An individual covariate selection for each marker-outcome-combination was outside the scope of this investigation and would also have compromised comparability of effect sizes between markers within this study. Future studies with a stronger focus on causal associations and the mechanistic relationship between markers and outcomes are needed to deepen our understanding. Thirdly, as with any study that reports on a large number of individual statistical tests, multiple testing is an issue. Here, we adjusted the p-values using the Bonferroni correction. While this correction is robust, it also comes with the risk of missing true effects due to its conservative approach. Therefore, it is possible that true associations did not reach statistical significance in our study. Finally, the comparatively small age range in BASE-II might be a reason for the weak correlation between some markers and chronological age and limit our conclusions to this age group. Future longitudinal studies with a wider age range and repeated marker measurements are needed to further clarify our understanding of longitudinal associations between the markers investigated here and clinical aging phenotypes.

Strengths of these analyses include the comparative analysis of a large number of markers derived from a wide variety of aging domains including epigenetics, proteomics, telomeres, composite markers, psychological markers, SkinAge, and BrainAge. The availability of these variables in addition to numerous age-associated outcomes in a large longitudinal sample allowed a comprehensive comparison of these markers cross-sectionally as well as over time. We provide information on the distinct abilities of these markers which can inform future studies and sharpen their profiles. As the measurement of numerous markers is costly, these analyses allow a targeted selection of markers that can be used in future studies based on their strength of association with the specific ageing domain of interest.

## Conclusion

Our comparative analyses of 16 markers of aging in the context of a wide range of age-associated outcome variables highlight the distinction between the investigated domains of aging. Interestingly, markers were more frequently and also more strongly associated with outcomes on average 7.4 years after their assessment compared to cross-sectional analyses suggesting their sensitivity to biological aging processes that became clinically apparent only much later during the follow-up examination. This finding underscores the potential of these markers for early risk stratification. Namely, ALI and DunedinPACE substantially contributed to the prediction of incident impairment at follow-up examination.

## Acknowledgments

Funding

This work was supported by grants of the Deutsche Forschungsgemeinschaft (project number 460683900 to ID and LB), the ERC (as part of the “Lifebrain” project to LB and CML), and the Cure Alzheimer’s Fund (as part of the “CIR-CUITS” consortium to LB), and a grant from the EU Joint Programme – Neurodegenerative Disease Research (JPND2021-650-289, coordinator: CML). This article uses data from the Berlin Aging Study II (BASE-II). BASE-II was supported by the German Federal Ministry of Education and Research under grant numbers #01UW0808; #16SV5536K, #16SV5537, #16SV5538, #16SV5837, #01GL1716A, and #01GL1716B. C.M. L. was supported by the Heisenberg program of the DFG (DFG; LI 2654/4-1).

## Conflict of interest

None

## Data Availability

Due to concerns for participant privacy, data are available only upon reasonable request. Please contact Ludmila Müller, scientific coordinator, at, for additional information.

## Ethics

All participants gave written informed consent. The Ethics Committee of the Charité – Universitätsmedizin Berlin approved the study (approval numbers EA2/029/09 and EA2/144/16). The study was conducted in accordance with the Declaration of Helsinki and was registered in the German Clinical Trials Registry as DRKS00009277.

## Author contributions

Conceptualization: V.M.V, I.D.; Data curation: V.M.V., J.H., J. D., S.D., L.D., P.J., E.L., S.M.,; Formal analysis: V.M.V; Funding acquisition: E.S.-T., U.L., D.G., L.B., I.D., C.M.L.; Investigation: V.M.V, I.D.; Methodology: V.M.V, I.D.; Project administration: V.M.V, I.D.; Resources: S.K., M.M., E.S.-T., M.R., U.L., C.M.L., D.G., L.B., I.D.; Supervision: I.D.; Visualization: V.M.V; Writing - original draft: V.M.V, I.D.; and Writing - review & editing: all authors.

## Notes

### Competing Interest Statement

The authors have declared no competing interest.

### Author Declarations

The Ethics Committee of the Charite - Universitaetsmedizin Berlin gave ethical approval for the study (approval numbers EA2/029/09 and EA2/144/16).

## References

1. Oeppen J, Vaupel JW. Broken limits to life expectancy. American Association for the Advancement of Science; 2002. p. 1029–31.

2. Crimmins EM. Lifespan and healthspan: past, present, and promise. The Gerontologist. 2015;55(6):901–11.

3. Moqri M, Herzog C, Poganik JR, Justice J, Belsky DW, Higgins-Chen A, et al. Biomarkers of aging for the identification and evaluation of longevity interventions. Cell. 2023;186(18):3758–75.

4. Garmany A, Yamada S, Terzic A. Longevity leap: mind the healthspan gap. npj Regenerative Medicine. 2021;6(1):57.

5. Jylhava J, Pedersen NL, Hagg S. Biological Age Predictors. EBioMedicine. 2017;21:29–36.

6. Kennedy BK, Berger SL, Brunet A, Campisi J, Cuervo AM, Epel ES, et al. Geroscience: Linking Aging to Chronic Disease. Cell. 2014;159(4):709–13.

7. Goldman DP, Cutler D, Rowe JW, Michaud P-C, Sullivan J, Peneva D, et al. Substantial health and economic returns from delayed aging may warrant a new focus for medical research. Health affairs. 2013;32(10):1698–705.

8. Kirkland JL. Translating the science of aging into therapeutic interventions. Cold Spring Harbor perspectives in medicine. 2016;6(3):a025908.

9. Hägg S, Belsky DW, Cohen AA. Developments in molecular epidemiology of aging. Emerging Topics in Life Sciences. 2019;3(4):411–21.

10. Lara J, Cooper R, Nissan J, Ginty AT, Khaw K-T, Deary IJ, et al. A proposed panel of biomarkers of healthy ageing. BMC medicine. 2015;13(1):1–8.

11. Cummings SR, Kritchevsky SB. Endpoints for geroscience clinical trials: health outcomes, biomarkers, and biologic age. GeroScience. 2022;44(6):2925–31.

12. Macdonald-Dunlop E, Taba N, Klarić L, Frkatović A, Walker R, Hayward C, et al. A catalogue of omics biological ageing clocks reveals substantial commonality and associations with disease risk. Aging. 2022;14(2):623–59.

13. Raj K, Horvath S. Current perspectives on the cellular and molecular features of epigenetic ageing. Experimental Biology and Medicine. 2020;245(17):1532–42.

14. Belsky DW, Caspi A, Corcoran DL, Sugden K, Poulton R, Arseneault L, et al. DunedinPACE, a DNA methylation biomarker of the pace of aging. eLife. 2022;11:e73420.

15. Drewelies J, Hueluer G, Duezel S, Vetter VM, Pawelec G, Steinhagen-Thiessen E, et al. Using blood test parameters to define biological age among older adults: association with morbidity and mortality independent of chronological age validated in two separate birth cohorts. GeroScience. 2022.

16. Seeman TE, Singer BH, Rowe JW, Horwitz RI, McEwen BS. Price of adaptation— allostatic load and its health consequences: MacArthur studies of successful aging. Archives of internal medicine. 1997;157(19):2259–68.

17. Belsky DW, Moffitt TE, Cohen AA, Corcoran DL, Levine ME, Prinz JA, et al. Eleven telomere, epigenetic clock, and biomarker-composite quantifications of biological aging: do they measure the same thing? American journal of epidemiology. 2018;187(6):1220–30.

18. Vetter VM, Meyer A, Karbasiyan M, Steinhagen-Thiessen E, Hopfenmuller W, Demuth I. Epigenetic clock and relative telomere length represent largely different aspects of aging in the Berlin Aging Study II (BASE-II). The journals of gerontology Series A, Biological sciences and medical sciences. 2018.

19. Vetter VM, Kalies CH, Sommerer Y, Spira D, Drewelies J, Regitz-Zagrosek V, et al. Relationship Between 5 Epigenetic Clocks, Telomere Length, and Functional Capacity Assessed in Older Adults: Cross-Sectional and Longitudinal Analyses. The Journals of Gerontology: Series A. 2022;77(9):1724–33.

20. McCrory C, Fiorito G, McLoughlin S, Polidoro S, Cheallaigh CN, Bourke N, et al. Epigenetic clocks and allostatic load reveal potential sex-specific drivers of biological aging. The Journals of Gerontology: Series A. 2020;75(3):495–503.

21. Kim S, Myers L, Wyckoff J, Cherry KE, Jazwinski SM. The frailty index outperforms DNA methylation age and its derivatives as an indicator of biological age. GeroScience. 2017;39(1):83–92.

22. Zhang Y, Saum K-U, Schöttker B, Holleczek B, Brenner H. Methylomic survival predictors, frailty, and mortality. Aging. 2018;10(3):339.

23. Li X, Ploner A, Wang Y, Magnusson PK, Reynolds C, Finkel D, et al. Longitudinal trajectories, correlations and mortality associations of nine biological ages across 20-years follow-up. Elife. 2020;9:e51507.

24. Drewelies J, Homann J, Vetter V, Duezel S, Kühn S, Deecke L, et al. There are multiple clocks that time us: Cross-sectional and longitudinal associations among 14 alternative indicators of age and aging. 2023.

25. Kuiper LM, Polinder-Bos HA, Bizzarri D, Vojinovic D, Vallerga CL, Beekman M, et al. Epigenetic and metabolomic biomarkers for biological age: a comparative analysis of mortality and frailty risk. The Journals of Gerontology: Series A. 2023;78(10):1753–62.

26. Bertram L, Bockenhoff A, Demuth I, Duzel S, Eckardt R, Li SC, et al. Cohort profile: The Berlin Aging Study II (BASE-II). International journal of epidemiology. 2014;43(3):703–12.

27. Demuth I, Banszerus V, Drewelies J, Düzel S, Seeland U, Spira D, et al. Cohort profile: follow-up of a Berlin Aging Study II (BASE-II) subsample as part of the GendAge study. BMJ Open. 2021;11(6):e045576.

28. von Elm E, Altman DG, Egger M, Pocock SJ, Gøtzsche PC, Vandenbroucke JP. The Strengthening the Reporting of Observational Studies in Epidemiology (STROBE) statement: guidelines for reporting observational studies. Lancet. 2007;370(9596):1453-7.

29. Horvath S. DNA methylation age of human tissues and cell types. Genome biology. 2013;14(10):R115.

30. Hannum G, Guinney J, Zhao L, Zhang L, Hughes G, Sadda S, et al. Genome-wide methylation profiles reveal quantitative views of human aging rates. Molecular cell. 2013;49(2):359–67.

31. Levine ME, Lu AT, Quach A, Chen BH, Assimes TL, Bandinelli S, et al. An epigenetic biomarker of aging for lifespan and healthspan. Aging. 2018;10(4):573.

32. Lu AT, Quach A, Wilson JG, Reiner AP, Aviv A, Raj K, et al. DNA methylation GrimAge strongly predicts lifespan and healthspan. Aging. 2019;11(2):303.

33. Vetter VM, Kalies CH, Sommerer Y, Bertram L, Demuth I. Seven-CpG DNA Methylation Age Determined by Single Nucleotide Primer Extension and Illumina’s Infinium MethylationEPIC Array Provide Highly Comparable Results. Frontiers in genetics. 2022;12.

34. Meyer A, Salewsky B, Buchmann N, Steinhagen-Thiessen E, Demuth I. Relative Leukocyte Telomere Length, Hematological Parameters and Anemia - Data from the Berlin Aging Study II (BASE-II). Gerontology. 2016;62(3):330–6.

35. Lu AT, Seeboth A, Tsai P-C, Sun D, Quach A, Reiner AP, et al. DNA methylation-based estimator of telomere length. Aging. 2019;11(16):5895.

36. Seeman M, Merkin SS, Karlamangla A, Koretz B, Seeman T. Social status and biological dysregulation: The “status syndrome” and allostatic load. Social Science & Medicine. 2014;118:143–51.

37. McCrory C, Fiorito G, Cheallaigh CN, Polidoro S, Karisola P, Alenius H, et al. How does socio-economic position (SEP) get biologically embedded? A comparison of allostatic load and the epigenetic clock (s). Psychoneuroendocrinology. 2019;104:64–73.

38. Rubin DC, Berntsen D. People over forty feel 20% younger than their age: Subjective age across the lifespan. Psychonomic bulletin & review. 2006;13(5):776–80.

39. Düzel S, Voelkle MC, Düzel E, Gerstorf D, Drewelies J, Steinhagen-Thiessen E, et al. The Subjective Health Horizon Questionnaire (SHH-Q): assessing future time perspectives for facets of an active lifestyle. Gerontology. 2016;62(3):345–53.

40. Jawinski P, Markett S, Drewelies J, Düzel S, Demuth I, Steinhagen-Thiessen E, et al. Linking brain age gap to mental and physical health in the Berlin aging study II. Frontiers in Aging Neuroscience. 2022;14:791222.

41. Lemke E, Vetter VM, Berger N, Banszerus VL, König M, Demuth I. Cardiovascular health is associated with the epigenetic clock in the Berlin Aging Study II (BASE-II). Mechanisms of ageing and development. 2022;201:111616.

42. Vetter VM, Demircan K, Homann J, Chillon TS, Muelleder M, Shomroni O, et al. Low Blood Levels of Selenium, Selenoprotein P and GPx3 are Associated with Accelerated Biological Aging: Results from the Berlin Aging Study II (BASE-II). medRxiv. 2024:2024.04. 04.24305314.

43. Vetter VM, Drewelies J, Sommerer Y, Kalies CH, Regitz-Zagrosek V, Bertram L, et al. Epigenetic aging and perceived psychological stress in old age. Translational Psychiatry. 2022;12(1):410.

44. Vetter VM, Sommerer Y, Kalies CH, Spira D, Bertram L, Demuth I. Vitamin D supplementation is associated with slower epigenetic aging. GeroScience. 2022.

45. Vetter VM, Spieker J, Sommerer Y, Buchmann N, Kalies CH, Regitz-Zagrosek V, et al. DNA methylation age acceleration is associated with risk of diabetes complications. Communications Medicine. 2023;3(1):21.

46. Vetter VM, Spira D, Banszerus VL, Demuth I. Epigenetic clock and leukocyte telomere length are associated with vitamin D status, but not with functional assessments and frailty in the Berlin Aging Study II. The Journals of Gerontology: Series A. 2020.

47. Schmid H, Vetter VM, Homann J, Bahr V, Lill CM, Regitz-Zagrosek V, et al. Cross-sectional and Longitudinal Relationship between Sex Hormones and Six Epigenetic Clocks in Older Adults: Results of the Berlin Aging Study II (BASE-II). medRxiv. 2024:2024.11. 15.24317371.

48. Demuth I, Vetter VM, Homann J, Regitz-Zagrosek V, Gerstorf D, Lill CM, et al. DunedinPACE Predicts Incident Metabolic Syndrome: Cross-sectional and Longitudinal Data from the Berlin Aging Study II (BASE-II). medRxiv. 2024:2024.12. 16.24319083.

49. Drewelies J, Homann J, Vetter VM, Duezel S, Kühn S, Deecke L, et al. There are multiple clocks that time us: Cross-sectional and longitudinal associations among 14 alternative indicators of age and aging. The Journals of Gerontology, Series A: Biological Sciences and Medical Sciences. 2024:glae244.

50. Fried LP, Tangen CM, Walston J, Newman AB, Hirsch C, Gottdiener J, et al. Frailty in older adults: evidence for a phenotype. The Journals of Gerontology Series A: Biological Sciences and Medical Sciences. 2001;56(3):M146–M57.

51. Vetter VM, Drewelies J, Düzel S, Homann J, Meyer-Arndt L, Braun J, et al. Change in body weight of older adults before and during the COVID-19 pandemic: Longitudinal results from the Berlin Aging Study II. The Journal of nutrition, health and aging. 2024;28(4):100206.

52. Rockwood K, Mitnitski A. Frailty in relation to the accumulation of deficits. The journals of gerontology Series A, Biological sciences and medical sciences. 2007;62(7):722–7.

53. Folstein MF, Folstein SE, McHugh PR. “Mini-mental state”. A practical method for grading the cognitive state of patients for the clinician. Journal of psychiatric research. 1975;12(3):189–98.

54. 54. San Antonio TPC. Wechsler Adult Intelligence Scale - Revised. 1981.

55. Radloff LS. The CES-D Scale:A Self-Report Depression Scale for Research in the General Population. Applied Psychological Measurement. 1977;1(3):385–401.

56. Mahoney FI, Barthel DW. FUNCTIONAL EVALUATION: THE BARTHEL INDEX. Maryland state medical journal. 1965;14:61–5.

57. Vellas B, Guigoz Y, Garry PJ, Nourhashemi F, Bennahum D, Lauque S, et al. The Mini Nutritional Assessment (MNA) and its use in grading the nutritional state of elderly patients. Nutrition. 1999;15(2):116–22.

58. Association AD. 2. Classification and diagnosis of diabetes: standards of medical care in diabetes—2019. Diabetes care. 2019;42(Supplement 1):S13–S28.

59. Young BA, Lin E, Von Korff M, Simon G, Ciechanowski P, Ludman EJ, et al. Diabetes complications severity index and risk of mortality, hospitalization, and healthcare utilization. The American journal of managed care. 2008;14(1):15.

60. Spieker J, Vetter VM, Drewelies J, Spira D, Steinhagen-Thiessen E, Regitz-Zagrosek V, et al. Diabetes type 2 in the Berlin Aging Study II: Cross-sectional and longitudinal data on prevalence, incidence and severity over on average seven years of follow-up. Diabetic Medicine. 2023;n/a(n/a):e15104.

61. Charlson ME, Pompei P, Ales KL, MacKenzie CR. A new method of classifying prognostic comorbidity in longitudinal studies: development and validation. Journal of chronic diseases. 1987;40(5):373–83.

62. 62. Collaboration ECR, group Sw. SCORE2 risk prediction algorithms: new models to estimate 10-year risk of cardiovascular disease in Europe. European Heart Journal. 2021;42(25):2439–54.

63. 63. SCORE2-OP risk prediction algorithms: estimating incident cardiovascular event risk in older persons in four geographical risk regions. European Heart Journal. 2021;42(25):2455–67.

64. König M, Drewelies J, Norman K, Spira D, Buchmann N, Hülür G, et al. Historical trends in modifiable indicators of cardiovascular health and self-rated health among older adults: Cohort differences over 20 years between the Berlin Aging Study (BASE) and the Berlin Aging Study II (BASE-II). PloS one. 2018;13(1):e0191699.

65. Lloyd-Jones DM, Hong Y, Labarthe D, Mozaffarian D, Appel LJ, Van Horn L, et al. Defining and setting national goals for cardiovascular health promotion and disease reduction: the American Heart Association’s strategic Impact Goal through 2020 and beyond. Circulation. 2010;121(4):586–613.

66. Alberti KG, Eckel RH, Grundy SM, Zimmet PZ, Cleeman JI, Donato KA, et al. Harmonizing the metabolic syndrome: a joint interim statement of the International Diabetes Federation Task Force on Epidemiology and Prevention; National Heart, Lung, and Blood Institute; American Heart Association; World Heart Federation; International Atherosclerosis Society; and International Association for the Study of Obesity. Circulation. 2009;120(16):1640–5.

67. Hong S, Dobricic V, Ohlei O, Bos I, Vos SJB, Prokopenko D, et al. TMEM106B and CPOX are genetic determinants of cerebrospinal fluid Alzheimer’s disease biomarker levels. Alzheimers Dement. 2021;17(10):1628–40.

68. 68. Team RC. R: A language and environment for statistical computing. R Foundation for Statistical Computing, Vienna, Austria URL https://wwwR-projectorg. 2022.

69. Moqri M, Herzog C, Poganik JR, Ying K, Justice JN, Belsky DW, et al. Validation of biomarkers of aging. Nature medicine. 2024.

70. Hillary RF, Stevenson AJ, McCartney DL, Campbell A, Walker RM, Howard DM, et al. Epigenetic measures of ageing predict the prevalence and incidence of leading causes of death and disease burden. Clinical epigenetics. 2020;12(1):115.

71. DeLong ER, DeLong DM, Clarke-Pearson DL. Comparing the areas under two or more correlated receiver operating characteristic curves: a nonparametric approach. Biometrics. 1988:837–45.

