## Supplementary Material for "Comprehensive Comparison of Sixteen Markers of Biological Aging: Cross-Sectional and Longitudinal Results from the Berlin Aging Study II (BASE-II)"

Supplementary Methods

Biomarkers of Aging

DNA Methylation Age Acceleration (DNAmAA)

DNA methylation age (DNAmA) was estimated using the 7-CpG (1), Horvath (2), Hannum (3), PhenoAge (4), GrimAge (5), and DunedinPACE (6) clock algorithms. EDTA whole blood samples were drawn and stored at -80°C. DNA was isolated with the "Plus XL manual kit" (LGC). The 7-CpG clock was measured by Single Nucleotide Prime Extension (SNuPE) in 1,471 BASE-II samples. A detailed laboratory protocol was published before (1). The other clocks are calculated from methylation data measured with the "Infinium MethylationEPIC" array, version 1 (Illumina, Inc., USA) which is available for n=1,030 BASE-II participants. Data was loaded and processed with R's "Bigmelon" package (7). Outliers (outlyx function, myP = 0.15) and samples with insufficient bisulfite conversion efficacy (<80%) were removed and values were normalized (dasen function). The qual function was used to determine the extent of change between raw and normalized beta values. A root-mean-square deviation of >=0.1 led to the exclusion of the sample and loading and normalization was repeated with the updated sample. Finally, the raw and not normalized values were uploaded to Steve Horvath's website (https://horvath.genetics.ucla.edu/html/dnamage/) according to the manual. Additional information on the measurement of the epigenetic clocks in BASE-II can be found here (8). The DunedinPACE clock was calculated based on the instructions in the original publication by Belsky and colleagues (6).

DNAmAA was calculated for all clocks as residuals of a linear regression of DNAmA on chronological age, and leukocyte cell distribution (neutrophils, monocytes, lymphocytes, and eosinophils in G/l). Accelerated epigenetic age is indicated by positive DNAmAA values. As the DunedinPACE clock reflects the Pace of Aging, no DNAmAA for the DunedinPACE clock was calculated.

Proteomics Clock

Serum samples were semi-automatically processed in 96-well plates, including denaturation, reduction, alkylation, trypsin digestion, and cleanup steps (9). Liquid chromatography-mass spectrometry (LC-MS) analysis was conducted using a Bruker timsTOF Pro system coupled with an Agilent 1290 Infinity II LC system (10). Subsequently, a spectral library based on the Human Plasma Peptide Atlas and annotation of peptide sequences to the Uniprot human reference proteome (11, 12) was generated. Data annotation and quantifications was done with the software DIA-NN (13) and pre-processing was performed using MS-DAP (14) as framework and included data normalization, outlier sample filtering, low-presence peptide filtering, data imputation, and batch correction to ensure high data quality. Missing protein values were imputed using the knn method (15) employing the R package impute (16). Additional details on measurement and quality control can be found in ref. (17). 248 proteins were available for the training of the proteomics clock in BASE-II. In a first step, a 20-fold cross-validation (3 repetitions) of a linear regression, ridge regression, lasso regression and elastic-net regression of chronological age on all available proteins was calculated to assess model performance. Cross-validation was done with R's caret package (18). $R^2$, Mean Absolute Error (MAE) and Root Mean Squared Error (RMSE) were used to assess model performance. Best results were achieved using the elastic-net model (fraction = 0.525, lambda = 0.1, RMSE=3.55, $R^2$ = 0.11, MAE = 2.85). For the training of the final proteomics clock, the whole data sample was used. Correlation between proteomics clock and chronological age was moderate to strong (Pearson's r = 0.51). Analog to the approach used for the epigenetic clocks, proteomics age acceleration was calculated as unstandardized residuals of a linear regression of proteomics age on chronological age.

Telomere Length (TL)

Telomeres, the ends of chromosomes, shorten with every cell replication cycle and are an established aging indicator. In this study, TL was measured from two different methods. First, quantitative real-time PCR was used to determine the relative leukocyte TL (rLTL). A detailed description of the protocol and the subsequent rLTL calculation can be found in ref. (19). Secondly, TL was estimated from DNA methylation data by the recently published algorithm by Lu and colleagues (20). The DNA methylation data derived TL is named DNAmTL in this study.

SkinAge

Photographs of the participants hands were scored by three independent raters with respect to the number of lentigines on the participants skin (0 = “no or very few lentigines” to 3 = “very abundant presence of lentigines on both hands”). SkinAge is the unit-weighted average of the score documented by the three raters (21) and higher values indicate more lentigines. Additional information on this marker can be found in ref. (21).

BioAge

BioAge is a composite marker that combines 13 routine laboratory values that were select for their ability to predict mortality: zinc, sodium, chloride, uric acid, albumin, alpha-1 globulin, alpha-2 globulin, HbA1c, hemoglobin, leukocytes, lymphocytes, and creatinine. BioAge was developed with data from the BASE cohort which was assessed prior to the BASE-II and is entirely independent from the sample analyzed in this study. A detailed description on the methodological approach and the construction of this marker can be found in ref. (21).

Allostatic Load (ALI)

The Allostatic Load Index (ALI) was computed as "Group ALI", a count-based formula that was used in the publication by Seeman and colleagues in 1997 (22) and is the most used version of the ALI today (23). Based on the sex-stratified individual distribution of each biomarker, participants who fall in the high-risk quartile (>75th percentile or <25th percentile) gain one additional point in their ALI. The ALI used in this study includes six domains: neuroendocrine (cortisol, dehydroepiandrosterone sulfate (DHEA-S)), inflammation (C-reactive protein), metabolic (high-density lipoprotein cholesterol (HDL-C), low-density lipoprotein cholesterol (LDL-C), triglycerides (TG), glycosylates hemoglobin (HbA1c), fasting glucose), renal (creatinine), cardiovascular (systolic blood pressure (SBP), diastolic blood pressure (DBP), resting heart rate (RHR)), and anthropometric (waist-hip-ration (WHR), body mass index (BMI)).To identify dysregulation that is possibly masked through successful pharmacological therapy (24, 25) the medication documented for each participant was considered during the calculation of the ALI. Independent of the measured variables, participants that reported the intake of medication were scored as if they were in the high-risk quartile of the regarding ALI component. This was done for the variables LDL-C (statins, cholesterol absorption inhibitors, niacin or bile sequestrants), TG (fibrates), HbA1c and fasting glucose (antidiabetic medication), SBP (antihypertensive medication), and RHR (beta-blockers, calcium channel blockers, cardiac glycosides, amiodaron). The final ALI can assume values between 0 and 14. The contributing parameters and considered medications were chosen in accordance with the publications by McCrory et al. and Seeman et al. (22, 24, 25).

Subjective Felt Age (SFA)

Participants reported how old they feel in years. To adjust for chronological age and in line with previous research, proportional discrepancy scores were calculated by dividing the difference between chronological and SFA through chronological age (26). Participants with a proportional discrepancy score that differed more than three standard deviations from the mean were excluded from the analyses (n=5) (27).

Subjective Life Expectancy (SLE)

Participants were asked to state the age at which they expect to die as part of the Subjective Health Horizon Questionnaire (SHH-Q) questionnaire ("Please estimate how old you will approximately become:…", (28)). The numeric age in years was then subtracted from chronological age. Detailed information on SLE can be found in ref. (28, 29).

Subjective Health Expectancy (SHE)

Participants were asked to predict the age up to which they will be healthy ("Please assess up to which age you will feel healthy."). SHE was subsequently subtracted from chronological age. Detailed information on this indicator of aging can be found in ref. (28, 29).

BrainAge

In a subgroup of BASE-II participants (N=255), BrainAge was measured from Magnetic Resonance Imaging (MRI) obtained from a 3-Tesla Siemens Magnetom Trio scanner with an age estimation model trained on data from 32,634 participants of the UK Biobank (30). The BrainAge variable used here was residualized for sex, age, age^2, and Total Intracranial Volume.

Outcomes

Fried's Frailty Index

Fried's Frailty Index was calculated as described in the original publication (31) by aggregating information on unintended weight loss, exhaustion, weakness, slow walking speed, and low physical activity. Impairment, defined by pre-specified cut-off values, in each variable adds one point to the final index resulting in a possible range between 0 and 5 points. Cut-offs for impairment in each variable were chosen in accordance with the original publication by Fried and colleagues. Additional information on Fried's Frailty Index in BASE-II can be found in ref. (32).

SPRINT-BASEed Frailty Index

A modified version of the Frailty Index proposed by Pajewsky and colleagues (33) was calculated from 31 of the originally described 37 items in the BASE-II dataset. This Frailty Index employs a deficit accumulation approach and was developed from data of the Systolic Blood Pressure Intervention Trial (SPRINT). In addition to the variables proposed in the original manuscript, grip strength was included in the modified version of the frailty index resulting in a total of 32 items of which no less than 30 had to be available in each participant to be included in the final dataset. Further information about the SPRINT-BASEed Frailty Index are detailed in ref. (34).

Finger Floor Distance (FFD)

Participants were asked to bend at the waist and reach the floor with their fingertips while their legs remained outstretched and their feed stood together (35). The distance between fingertips and floor was measured in centimeters and is used to evaluate the participants' mobility.

Mini-Mental State Examination (MMSE)

The MMSE (36) is a short interviewer-administered instrument to measure and monitor cognitive impairment. It contains a combination of questions and easy tasks that are to be performed by the participant. The MMSE tests orientation, attention, learning, calculation, delayed recall, and construction (37). An unauthorized version of the German MMSE was used by the study team without permission and has been rectified with PAR. The MMSE is a copyrighted instrument and may not be used or reproduced in whole or in part, in any form or language, or by any means without written permission of PAR (www.parinc.com).

Digit Symbol Substitution Test (DSST)

The DSST (38) is a well-validated pen-and-pencil test for cognitive function. Participants are asked to match symbols to numbers on a single sheet of paper according to a specified key on the top of the same page. The correctly allocated symbols are subsequently counted and the results can be used to assess and monitor cognitive performance (39).

Center for Epidemiological Studies Depression Scale (CES-D)

The CES-D is a short and self-administered 20-item questionnaire to assess depressive symptoms. It is designed to be used in studies including participants from the general population (40). Among others, the questionnaire contains items about appetite, ability to concentrate, sleep, happiness as well as the feeling of loneliness.

Activities of Daily Living Index (ADL)

The Activities of Daily Living Index (also known as Barthel Index) (41) is a 10-item questionnaire that assesses the ability to perform daily tasks without help. The questionnaire contains, among others, questions about personal toilet, continence, dressing and eating.

Mini Nutritional Assessment (MNA)

The MNA was developed to rapidly assess the nutritional status in older patients in nursing homes, outpatient clinics, and hospitals (42). It consists of a short questionnaire as well as the measurement of the circumference of the upper arm and calf.

Type 2 Diabetes (T2D)

T2D was diagnosed if at least one of the following criteria was fulfilled: Anamnestic history of T2D (self-report), antidiabetic medication, fasting plasma glucose ≥126 mg/dL, 2h plasma glucose during 75 g-OGTT ≥200 mg/dL, and HbA1c ≥ 48 mmol/mol [6.5%]. This approach is based on the American Diabetes Association (ADA) guidelines (43). Additional information about the diagnosis of T2D in BASE-II is provided in ref. (44).

Diabetes Complications Severity Index (DCSI)

The DCSI by Young and colleagues (45) is an instrument designed to quantify the severity of diabetes-associated complications and combines information on retinopathy, nephropathy, neuropathy, cerebrovascular disease, cardiovascular disease, peripheral vascular disease, and metabolic complications. Severity of each of the included complications was rated according to the original manuscript on a scale between 0 (=”no abnormality”) and 2 (=”severe abnormality”). In this study, a slightly adapted version of the DCSI is used since no information on the metabolic status was available (44).

Morbidity Index (MI)

A modified version of the Charlson Morbidity Index (46) was calculated for BASE-II participants to get an overall estimation of the participant’s morbidity (47). It includes information on myocardial infarction, congestive heart failure, peripheral vascular disease, cerebrovascular disease, dementia, chronic pulmonary disease, connective tissue disease, mild liver disease, diabetes, diabetes with end-organ damage, renal disease, hemiplegia, lymphoma, leukemia, any tumor, and moderate to severe liver disease.

SCORE2 and SCORE2-OP

SCORE2 (48) and SCORE2-OP (for participants >70 years, (49)) was calculated according to the procedure in the original publication based on age, smoking status, systolic blood pressure, and total- and HDL-cholesterol. As recommended by the author of the original publication, the score was not calculated for participants with diagnosed diabetes mellitus, myocardial infarction, or stroke. However, missing values for SCORE2/SCORE2-OP were still imputed to not loose these participants for the comparison analyses. We acknowledge that this procedure contrasts with the recommendations of the authors of the original publication but believe that SCORE2/SCORE2-OP provides valuable information about cardiovascular health even if it cannot be used for clinical decision making for patients diagnosed with diabetes mellitus, myocardial infarction, or stroke. For simplicity, hereafter SCORE2 and SCORE2-OP are both referred to as SCORE2. The same cut-off for impairment (>10%) was used both for the SCORE2 and SCORE2-OP.

Life's Simple 7 (LS7)

A slightly adapted version (50) of the Life's Simple 7 developed by the American Heart Association (AHA) (51) was calculated to assess the overall cardiovascular health of the participants based on modifiable cardiovascular risk factors. The score was built from data on BMI, blood pressure, total cholesterol, HbA1c, diet, smoking and physical activity as described before (50, 51).

Metabolic Syndrome (MetS)

The definition of the American Heart Association/ International Diabetes Federation/ National Heart, Lung, and Blood Institute criteria 2009 (52) was used to diagnose metabolic syndrome in BASE-II participants. Participants with values above the respective cut-off in at least three of the five criteria (waist circumference, triglycerides, HDL-cholesterol, blood pressure and fasting glucose) were diagnosed with metabolic syndrome. As described in the original publication, intake of respective medication also qualified for the diagnosis.

### Covariates

To allow comparison of effect sizes between investigated biomarkers, the same set of covariates was used for all analyses. Chronological age in years was calculated from the date of birth provided by the participants during a one-to-one interview with trained study personnel. Alcohol consumption was assessed in gram per day from a food frequency questionnaire (53). Smoking was recorded in packyears and BMI was calculated from bodyweight and height ($kg/m^2$) measured with the 763 seca measuring station (SECA, Germany). Genetic ancestry was assessed as the first four components of a principal component analysis on genome-wide SNP genotyping data (54).

### Missing Values

In the complete dataset including all independent and dependent variables and covariates, 6.5% of all values were missing and were imputed under the missing at random assumption with R's mice function (mice package (55), m = 5, maxit = 5, method = "pmm"). Distribution of the imputed variables was visually inspected and compared to the not-imputed variables distribution. All outcome variables at baseline and follow-up, covariates at baseline as well as all indicators of aging at baseline were included for the multiple imputations.

BrainAge, which was available in a subgroup of 255 participants, was not included in the imputation of the main dataset. A separate imputation was done including only the n=255 participants with available BrainAge values and all other markers of aging, outcome variables as well as confounding variables. All analyses of the main dataset were then repeated in this separately imputed dataset with BrainAge as indepent variable to facilitate a direct comparison of results from BrainAge with the other biomarkers in the same group of people.

Loss to follow-up

As described above, only participants that were part of the baseline as well as the follow-up examination have been analyzed in this study. This was done to facilitate comparison of the biomarkers' performance cross-sectionally and longitudinally in the same participants but also potentially introduces survival bias. However, this is a problem in many if not all longitudinal studies and we do not expect it to substantially influence our findings since differences between participants that were followed-up and that dropped out are generally small (Supplementary Table 1).

Statistical Analysis

Statistical computations and figures were produced using the R software package, version 4.3.2 (56). Descriptive statistics were calculated with the base and tableone package (57). Logistic and linear regression analyses were calculated with the g*lm* and *lm* function. To allow comparison of effect sizes between biomarkers and outcomes on different scales, the imputed datasets were normalized prior to the regression analyses with the *umx_scale* function (umx package (59)). Radar plots were drawn with the *radarchart* function of the fmsb package (62). The statistical significance between logistic regression models was assessed via likelihood ratio test (*lrtes* function from lmtest package (63)).

To facilitate the evaluation of the association between biomarkers and outcomes in a setting close to clinical application, continuously scaled outcome variables were dichotomized to reflect the individuals' status of impairment ("impaired" vs. "not-impaired"). Wherever possible, pre-defined and validated cut-off values were used. For DSST no validated cut-off was available and participants in the lowest quartile (<25$^{th}$ percentile) were defined as "impaired". The respective method of dichotomization as well as the used cut-off values are presented in Supplementary Table 3 and descriptive statistics of the dichotomized version of originally continuously scaled variables can be found in Supplementary Table 4.

To ensure consistency within the imputed datasets, dichotomization of continuous variables was done after the imputation and separately for each imputed dataset. This causes differences in the number of impaired cases and consequently in the sample size of the dataset analyzed with respect to incident cases in the longitudinal analyses (prevalent cases at baseline are excluded from the analyses). MMSE and ADL were excluded from analyses investigating the dichotomized version of originally contiguously scaled variables since only very few cases of impairment were observed. The individual number of participants analyzed is indicated with each statistical test.

Figures:

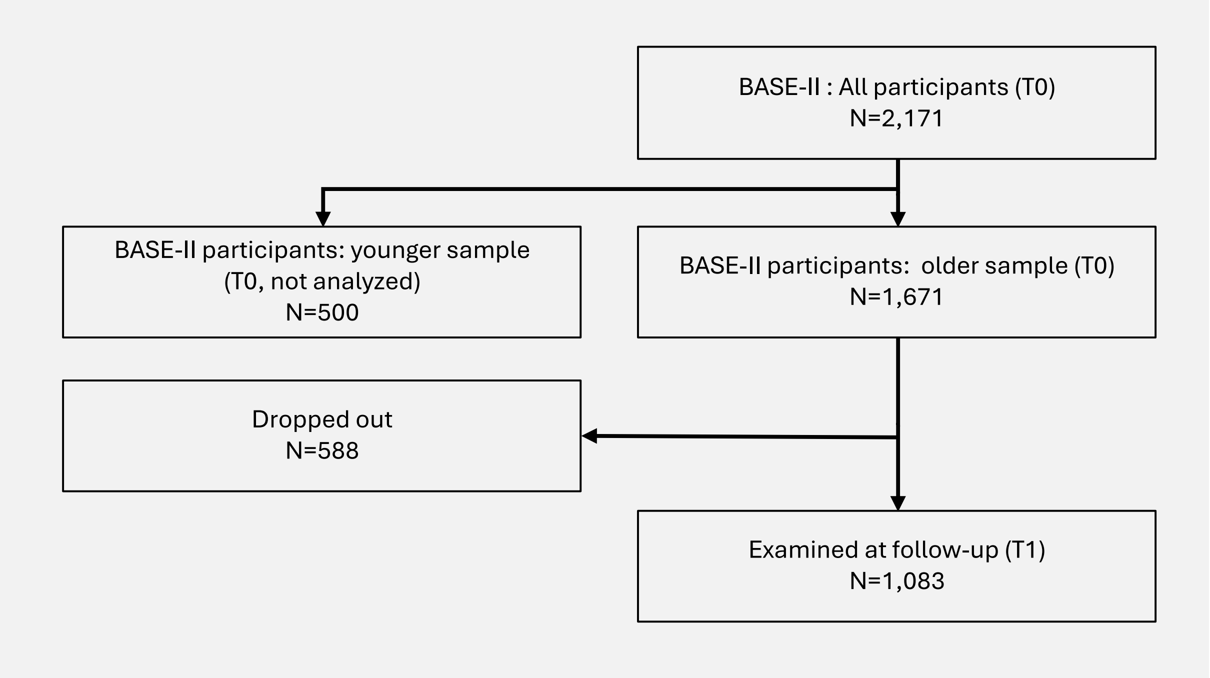


Supplementary Figure 1: Flowchart of participants of BASE-II that were analyzed in this study.

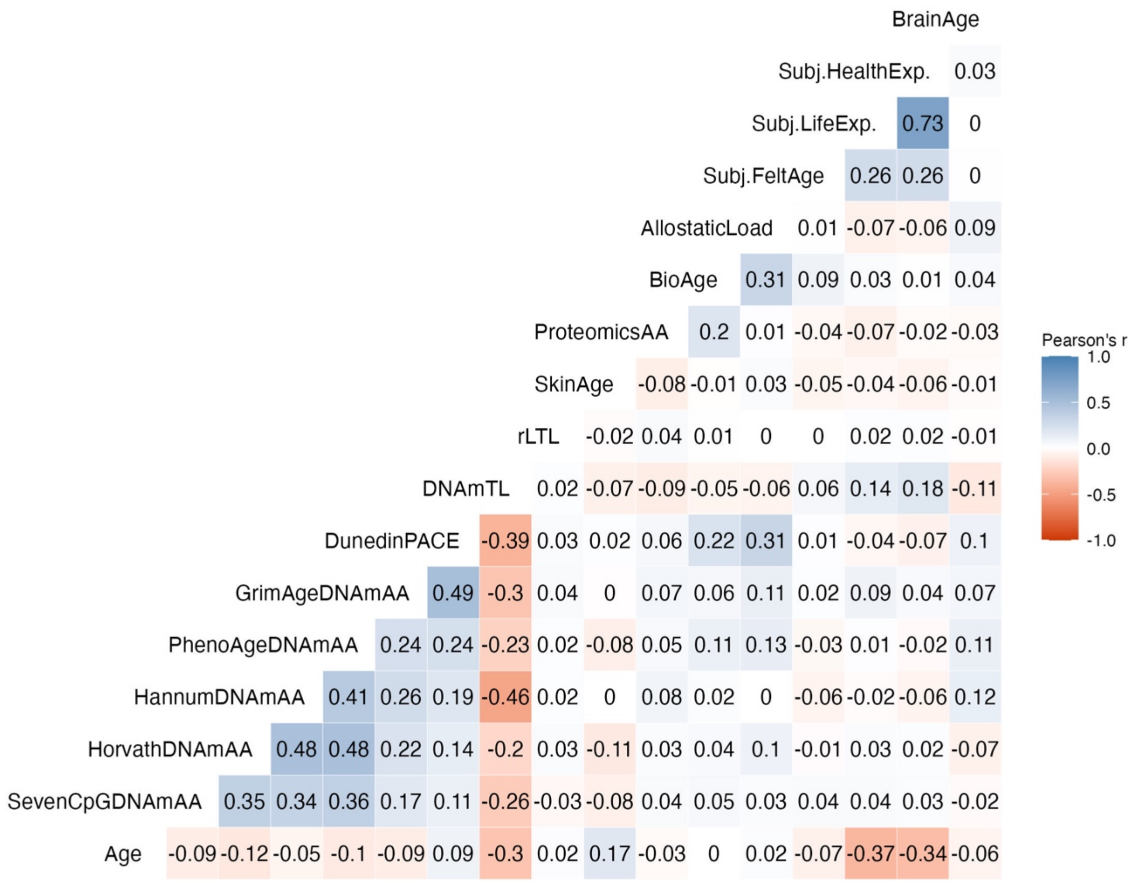


Supplementary Figure 2: Correlation plot of biomarkers of aging in the first imputed dataset (n=1,083). BrainAge was available for a smaller subgroup of 255 participants only.

A

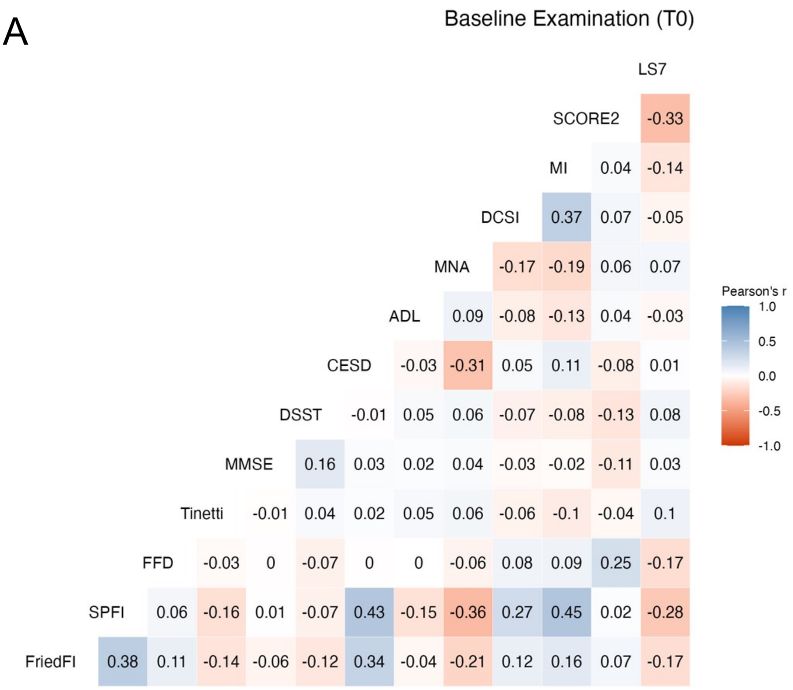


B

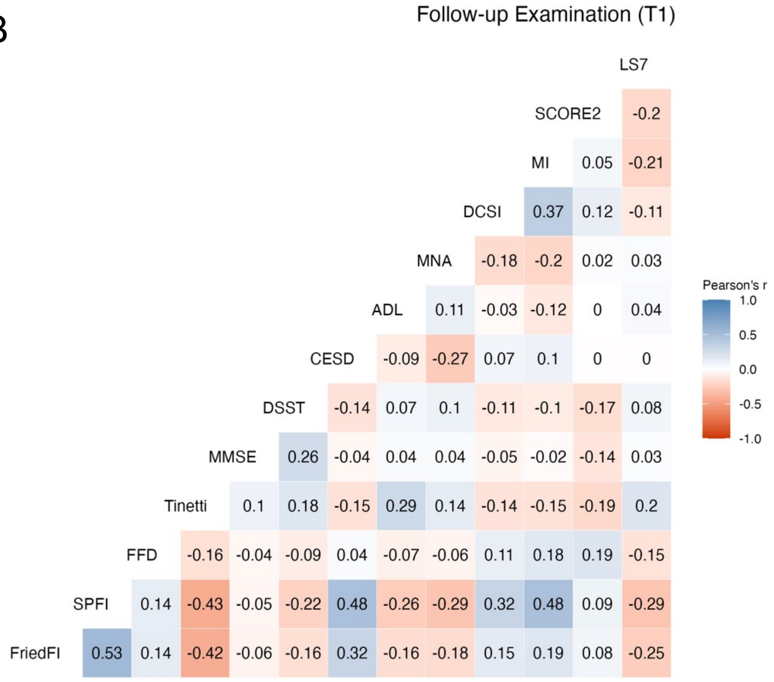


Supplementary Figure 3: Correlation plot of outcome variables at baseline (A) and at follow-up examination (B) in the first imputed dataset (n=1,083).
Note: LS7 = Life's Simple Seven, SCORE2 = Systematic Coronary Risk Evaluation 2, MI = Charlson's Morbidity Index, DCSI = Diabetes complications severity index, MNA = Mini Nutritional Assessment, DSST = Digit Symbol Substitution Test, Tinetti = Tinetti Test, FFD = Finger Floor Distance, SP FI = SPRINT-BASEed Frailty Phenotype, Fried FI = Fried's Frailty Phenotype, MetS = Metabolic Syndrome, T2DM = Type 2 Diabetes Mellitus.

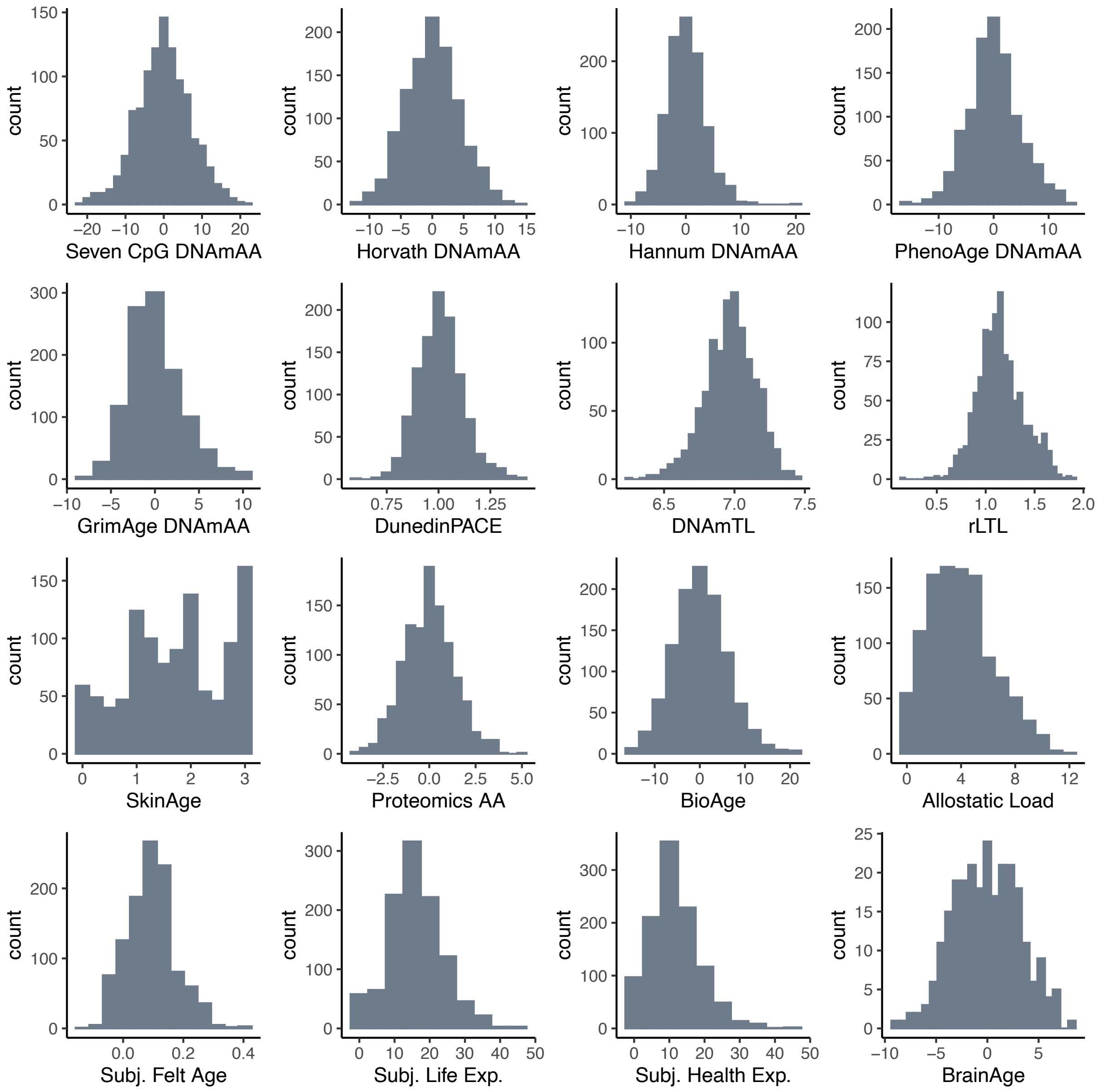


Supplementary Figure 4: Histograms of the biomarkers of aging in n=1,083 BASE-II participants. Information on BrainAge was available in a subgroup only (n=255).
Note: DNAmAA = DNA methylation age acceleration, DNAmTL = DNA methylation telomere length, rLTL = relative leukocyte telomere length, AA = age acceleration, Subj. = Subjective, Exp. = Expectancy.

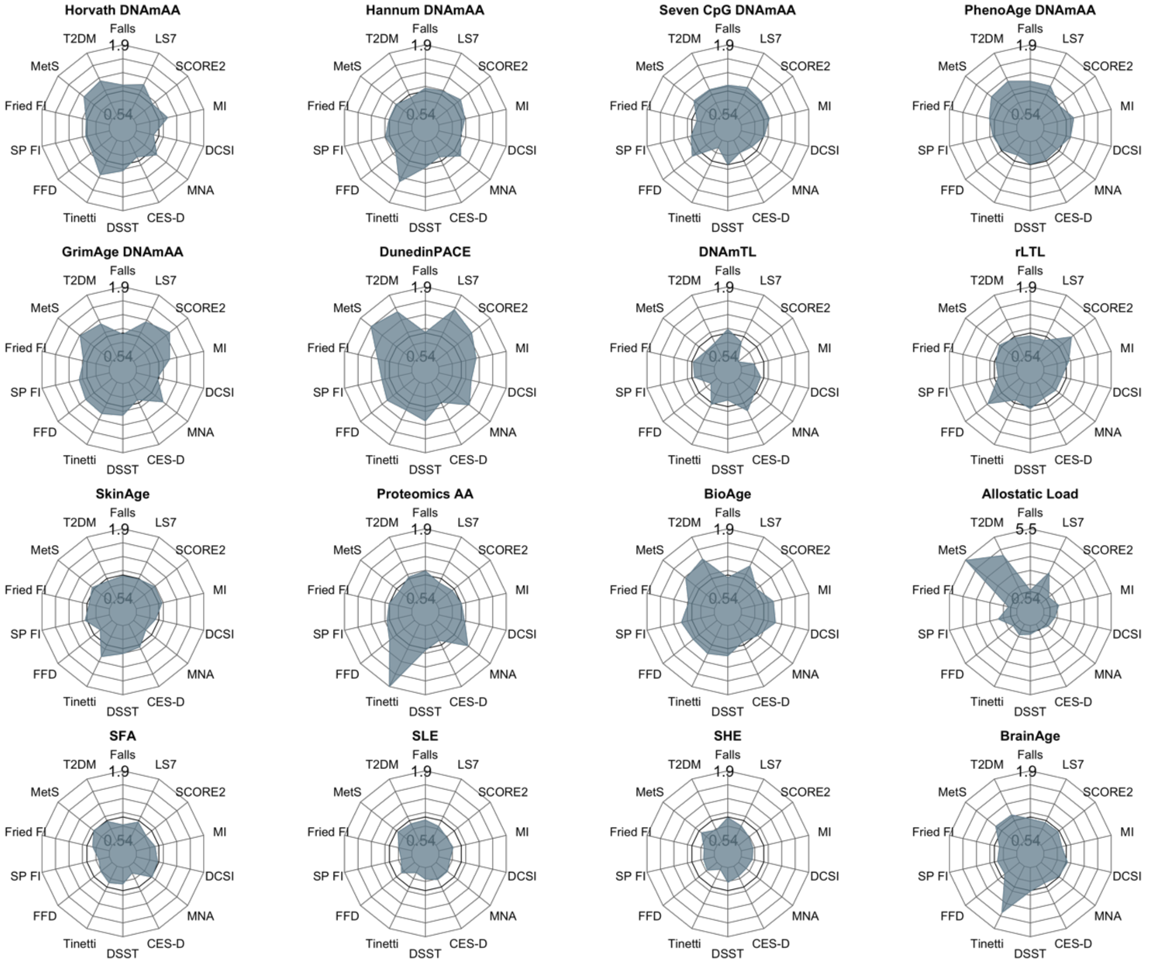


Supplementary Figure 5: Radar plot illustrating OR of logistic regression of binary and dichotomized outcome variables at T0 on biomarkers of aging at T0 in 1,083 BASE-II participants. All variables were standardized prior to the regression analysis. BrainAge was available for a smaller subgroup (n=255).

Note: LS7 = Life's Simple Seven, SCORE2 = Systematic Coronary Risk Evaluation 2, MI = Charlson's Morbidity Index, DCSI = Diabetes complications severity index, MNA = Mini Nutritional Assessment, DSST = Digit Symbol Substitution Test, Tinetti = Tinetti Test, FFD = Finger Floor Distance, SP FI = SPRINT-BASEed Frailty Phenotype, Fried FI = Fried's Frailty Phenotype, MetS = Metabolic Syndrome, T2DM = Type 2 Diabetes Mellitus.

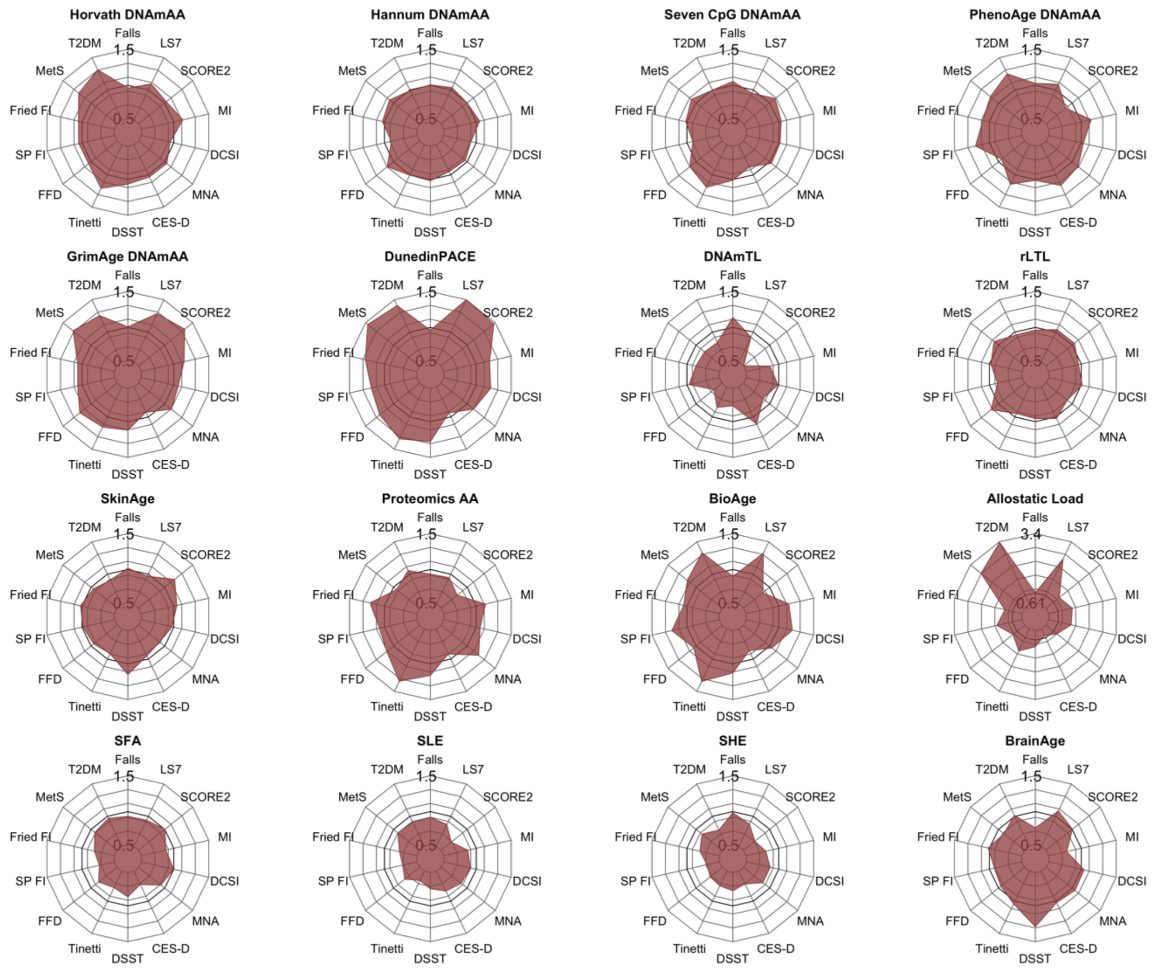


Supplementary Figure 6: Radar plot illustrating OR of logistic regression of binary and dichotomized outcome variables at T1 on biomarkers of aging at T0 in 1,083 BASE-II participants. All variables were standardized prior to the regression analysis. BrainAge was available for a smaller subgroup (n=255).

Note: LS7 = Life's Simple Seven, SCORE2 = Systematic Coronary Risk Evaluation 2, MI = Charlson's Morbidity Index, DCSI = Diabetes complications severity index, MNA = Mini Nutritional Assessment, DSST = Digit Symbol Substitution Test, Tinetti = Tinetti Test, FFD = Finger Floor Distance, SP FI = SPRINT-BASEed Frailty Phenotype, Fried FI = Fried's Frailty Phenotype, MetS = Metabolic Syndrome, T2DM = Type 2 Diabetes Mellitus.

References:


1. Vetter VM, Meyer A, Karbasiyan M, Steinhagen-Thiessen E, Hopfenmuller W, Demuth I. Epigenetic clock and relative telomere length represent largely different aspects of aging in the Berlin Aging Study II (BASE-II). The journals of gerontology Series A, Biological sciences and medical sciences. 2018.
2. Horvath S. DNA methylation age of human tissues and cell types. Genome biology. 2013;14(10):R115.
3. Hannum G, Guinney J, Zhao L, Zhang L, Hughes G, Sadda S, et al. Genome-wide methylation profiles reveal quantitative views of human aging rates. Molecular cell. 2013;49(2):359-67.
4. Levine ME, Lu AT, Quach A, Chen BH, Assimes TL, Bandinelli S, et al. An epigenetic biomarker of aging for lifespan and healthspan. Aging. 2018;10(4):573.
5. Lu AT, Quach A, Wilson JG, Reiner AP, Aviv A, Raj K, et al. DNA methylation GrimAge strongly predicts lifespan and healthspan. Aging. 2019;11(2):303.
6. Belsky DW, Caspi A, Corcoran DL, Sugden K, Poulton R, Arseneault L, et al. DunedinPACE, a DNA methylation biomarker of the pace of aging. eLife. 2022;11:e73420.
7. Gorrie-Stone TJ, Smart MC, Saffari A, Malki K, Hannon E, Burrage J, et al. Bigmelon: tools for analysing large DNA methylation datasets. Bioinformatics (Oxford, England). 2019;35(6):981-6.
8. Vetter VM, Spieker J, Sommerer Y, Buchmann N, Kalies CH, Regitz-Zagrosek V, et al. DNA methylation age acceleration is associated with risk of diabetes complications. Communications Medicine. 2023;3(1):21.
9. Messner CB, Demichev V, Wendisch D, Michalick L, White M, Freiwald A, et al. Ultra-high-throughput clinical proteomics reveals classifiers of COVID-19 infection. Cell systems. 2020;11(1):11-24. e4.
10. Szyrwiel L, Gille C, Mülleder M, Demichev V, Ralser M. Fast proteomics with dia-PASEF and analytical flow-rate chromatography. Proteomics. 2024;24(1-2):2300100.
11. UniProt: the universal protein knowledgebase in 2023. Nucleic acids research. 2023;51(D1):D523-D31.
12. Deutsch EW, Omenn GS, Sun Z, Maes M, Pernemalm M, Palaniappan KK, et al. Advances and utility of the human plasma proteome. Journal of proteome research. 2021;20(12):5241-63.
13. Demichev V, Messner CB, Vernardis SI, Lilley KS, Ralser M. DIA-NN: neural networks and interference correction enable deep proteome coverage in high throughput. Nature methods. 2020;17(1):41-4.
14. Koopmans F, Li KW, Klaassen RV, Smit AB. MS-DAP platform for downstream data analysis of label-free proteomics uncovers optimal workflows in benchmark data sets and increased sensitivity in analysis of Alzheimer's biomarker data. Journal of Proteome Research. 2022;22(2):374-86.
15. Troyanskaya O, Cantor M, Sherlock G, Brown P, Hastie T, Tibshirani R, et al. Missing value estimation methods for DNA microarrays. Bioinformatics (Oxford, England). 2001;17(6):520-5.
16. Hastie T, Tibshirani R, Narasimhan B, Chu G. impute: impute: Imputation for microarray data. R package version; 2020.
17. Dierks C, Mizrak RS, Shomroni O, Farztdinov V, Textoris-Taube K, Ludwig D, et al. Menopause Hormone Replacement Therapy and Lifestyle Factors affect Metabolism and Immune System in the Serum Proteome of Aging Individuals. medRxiv. 2024:2024.06. 22.24309293.
18. Kuhn M. Building predictive models in R using the caret package. Journal of statistical software. 2008;28:1-26.

19. Meyer A, Salewsky B, Buchmann N, Steinhagen-Thiessen E, Demuth I. Relative Leukocyte Telomere Length, Hematological Parameters and Anemia - Data from the Berlin Aging Study II (BASE-II). Gerontology. 2016;62(3):330-6.
20. Lu AT, Seeboth A, Tsai P-C, Sun D, Quach A, Reiner AP, et al. DNA methylation-based estimator of telomere length. Aging. 2019;11(16):5895.
21. Drewelies J, Hueluer G, Duezel S, Vetter VM, Pawelec G, Steinhagen-Thiessen E, et al. Using blood test parameters to define biological age among older adults: association with morbidity and mortality independent of chronological age validated in two separate birth cohorts. GeroScience. 2022.
22. Seeman TE, Singer BH, Rowe JW, Horwitz RI, McEwen BS. Price of adaptation—allostatic load and its health consequences: MacArthur studies of successful aging. Archives of internal medicine. 1997;157(19):2259-68.
23. Juster R-P, McEwen BS, Lupien SJ. Allostatic load biomarkers of chronic stress and impact on health and cognition. Neuroscience & Biobehavioral Reviews. 2010;35(1):2-16.
24. Seeman M, Merkin SS, Karlamangla A, Koretz B, Seeman T. Social status and biological dysregulation: The “status syndrome” and allostatic load. Social Science & Medicine. 2014;118:143-51.
25. McCrory C, Fiorito G, Cheallaigh CN, Polidoro S, Karisola P, Alenius H, et al. How does socio-economic position (SEP) get biologically embedded? A comparison of allostatic load and the epigenetic clock (s). Psychoneuroendocrinology. 2019;104:64-73.
26. Rubin DC, Berntsen D. People over forty feel 20% younger than their age: Subjective age across the lifespan. Psychonomic bulletin & review. 2006;13(5):776-80.
27. Weiss D, Lang FR. “They” are old but “I” feel younger: Age-group dissociation as a self-protective strategy in old age. Psychology and aging. 2012;27(1):153.
28. Düzel S, Voelkle MC, Düzel E, Gerstorf D, Drewelies J, Steinhagen-Thiessen E, et al. The Subjective Health Horizon Questionnaire (SHH-Q): assessing future time perspectives for facets of an active lifestyle. Gerontology. 2016;62(3):345-53.
29. Drewelies J, Homann J, Vetter V, Duezel S, Kühn S, Deecke L, et al. There are multiple clocks that time us: Cross-sectional and longitudinal associations among 14 alternative indicators of age and aging. 2023.
30. Jawinski P, Markett S, Drewelies J, Düzel S, Demuth I, Steinhagen-Thiessen E, et al. Linking brain age gap to mental and physical health in the Berlin aging study II. Frontiers in Aging Neuroscience. 2022;14:791222.
31. Fried LP, Tangen CM, Walston J, Newman AB, Hirsch C, Gottdiener J, et al. Frailty in older adults: evidence for a phenotype. The Journals of Gerontology Series A: Biological Sciences and Medical Sciences. 2001;56(3):M146-M57.
32. Spira D, Buchmann N, König M, Rosada A, Steinhagen-Thiessen E, Demuth I, et al. Sex-specific differences in the association of vitamin D with low lean mass and frailty–Results from the Berlin Aging Study II. Nutrition. 2018.
33. Rockwood K, Mitnitski A. Frailty in relation to the accumulation of deficits. The journals of gerontology Series A, Biological sciences and medical sciences. 2007;62(7):722-7.
34. Vetter VM, Drewelies J, Düzel S, Homann J, Meyer-Arndt L, Braun J, et al. Change in body weight of older adults before and during the COVID-19 pandemic: Longitudinal results from the Berlin Aging Study II. The Journal of nutrition, health and aging. 2024;28(4):100206.
35. Perret C, Poiraudeau S, Fermanian J, Colau MM, Benhamou MA, Revel M. Validity, reliability, and responsiveness of the fingertip-to-floor test. Archives of physical medicine and rehabilitation. 2001;82(11):1566-70.
36. Folstein MF, Folstein SE, McHugh PR. "Mini-mental state". A practical method for grading the cognitive state of patients for the clinician. Journal of psychiatric research. 1975;12(3):189-98.

37. Van Heugten CM, Walton L, Hentschel U. Can we forget the Mini-Mental State Examination? A systematic review of the validity of cognitive screening instruments within one month after stroke. Clinical Rehabilitation. 2015;29(7):694-704.
38. San Antonio TPC. Wechsler Adult Intelligence Scale - Revised. 1981.
39. Jaeger J. Digit Symbol Substitution Test: The Case for Sensitivity Over Specificity in Neuropsychological Testing. J Clin Psychopharmacol. 2018;38(5):513-9.
40. Radloff LS. The CES-D Scale:A Self-Report Depression Scale for Research in the General Population. Applied Psychological Measurement. 1977;1(3):385-401.
41. Mahoney FI, Barthel DW. FUNCTIONAL EVALUATION: THE BARTHEL INDEX. Maryland state medical journal. 1965;14:61-5.
42. Vellas B, Guigoz Y, Garry PJ, Nourhashemi F, Bennahum D, Lauque S, et al. The Mini Nutritional Assessment (MNA) and its use in grading the nutritional state of elderly patients. Nutrition. 1999;15(2):116-22.
43. Association AD. 2. Classification and diagnosis of diabetes: standards of medical care in diabetes—2019. Diabetes care. 2019;42(Supplement 1):S13-S28.
44. Spieker J, Vetter VM, Spira D, Steinhagen-Thiessen E, Regitz-Zagrosek V, Buchmann N, et al. Diabetes Type 2 in the Berlin Aging Study II: Prevalence, Incidence and Severity Over up to Ten Years of Follow-up. PREPRINT (Version 3) available at Research Square. 2021.
45. Young BA, Lin E, Von Korff M, Simon G, Ciechanowski P, Ludman EJ, et al. Diabetes complications severity index and risk of mortality, hospitalization, and healthcare utilization. The American journal of managed care. 2008;14(1):15.
46. Charlson ME, Pompei P, Ales KL, MacKenzie CR. A new method of classifying prognostic comorbidity in longitudinal studies: development and validation. Journal of chronic diseases. 1987;40(5):373-83.
47. Meyer A, Salewsky B, Spira D, Steinhagen-Thiessen E, Norman K, Demuth I. Leukocyte telomere length is related to appendicular lean mass: cross-sectional data from the Berlin Aging Study II (BASE-II). The American journal of clinical nutrition. 2016;103(1):178-83.
48. Collaboration ECR, group Sw. SCORE2 risk prediction algorithms: new models to estimate 10-year risk of cardiovascular disease in Europe. European Heart Journal. 2021;42(25):2439-54.
49. SCORE2-OP risk prediction algorithms: estimating incident cardiovascular event risk in older persons in four geographical risk regions. European Heart Journal. 2021;42(25):2455-67.
50. König M, Drewelies J, Norman K, Spira D, Buchmann N, Hülür G, et al. Historical trends in modifiable indicators of cardiovascular health and self-rated health among older adults: Cohort differences over 20 years between the Berlin Aging Study (BASE) and the Berlin Aging Study II (BASE-II). PloS one. 2018;13(1):e0191699.
51. Lloyd-Jones DM, Hong Y, Labarthe D, Mozaffarian D, Appel LJ, Van Horn L, et al. Defining and setting national goals for cardiovascular health promotion and disease reduction: the American Heart Association's strategic Impact Goal through 2020 and beyond. Circulation. 2010;121(4):586-613.
52. Alberti KG, Eckel RH, Grundy SM, Zimmet PZ, Cleeman JI, Donato KA, et al. Harmonizing the metabolic syndrome: a joint interim statement of the International Diabetes Federation Task Force on Epidemiology and Prevention; National Heart, Lung, and Blood Institute; American Heart Association; World Heart Federation; International Atherosclerosis Society; and International Association for the Study of Obesity. Circulation. 2009;120(16):1640-5.
53. Nöthlings U, Hoffmann K, Bergmann MM, Boeing HJTJon. Fitting portion sizes in a self-administered food frequency questionnaire. 2007;137(12):2781-6.

54. Hong S, Dobricic V, Ohlei O, Bos I, Vos SJB, Prokopenko D, et al. TMEM106B and CPOX are genetic determinants of cerebrospinal fluid Alzheimer's disease biomarker levels. Alzheimers Dement. 2021;17(10):1628-40.
55. van Buuren S, Groothuis-Oudshoorn K. mice: Multivariate Imputation by Chained Equations in R. Journal of Statistical Software. 2011;45(3):1 - 67.
56. Team RC. R: A language and environment for statistical computing. R Foundation for Statistical Computing, Vienna, Austria URL https://wwwR-projectorg. 2022.
57. Yoshida K. Bartel, A. tableone: Create "Table 1" to Describe Baseline Characteristics with or without Propensity Score Weights, version 0.13. 2, R Studio package; RStudio. Inc: Vienna, Austria. 2022.
58. Moqri M, Herzog C, Poganik JR, Ying K, Justice JN, Belsky DW, et al. Validation of biomarkers of aging. Nature medicine. 2024.
59. Bates TC, Maes H, Neale MC. umx: Twin and path-based structural equation modeling in R. Twin Research and Human Genetics. 2019;22(1):27-41.
60. Cohen J. Statistical power analysis for the behavioral sciences: routledge; 2013.
61. Fey CF, Hu T, Delios A. The measurement and communication of effect sizes in management research. Management and Organization Review. 2023;19(1):176-97.
62. Nakazawa M. fmsb: Functions for medical statistics book with some demographic data. R package version 06. 2018;3(3).
63. Zeileis A, Hothorn T. Diagnostic checking in regression relationships. 2002.
